# SHARED GENETIC ARCHITECTURE AND DIRECTIONAL ASSOCIATIONS BETWEEN ATTENTION-DEFICIT/HYPERACTIVITY AND BORDERLINE PERSONALITY DISORDERS: AN INTEGRATIVE GENOMIC ANALYSIS

**DOI:** 10.64898/2026.11.06.26367465

**Authors:** Gaetano Gorgone, Chiara Galati, Sofia Ciappina

## Abstract

**Background:** Attention-deficit/hyperactivity disorder (ADHD) and borderline personality disorder (BPD) frequently co-occur, but their shared genetic susceptibility and directional associations remain unclear.

**Methods:** We analyzed European-ancestry genome-wide association study (GWAS) data for ADHD (38,691 cases; 186,843 controls) and BPD (12,339 cases; 1,041,717 controls). Linkage disequilibrium score regression, conjunctional false discovery rate (conjFDR), bidirectional Mendelian randomization (MR), MR accounting for linkage disequilibrium and sample overlap (MR-LAP), Causal Analysis Using Summary Effect estimates (CAUSE), colocalization, and Sum of Single Effects (SuSiE) fine-mapping assessed genetic sharing and directionality.

**Results:** Genetic correlation was substantial (r_g = 0.733). ConjFDR identified 48 shared loci; 36 (75%) showed concordant directions in both phenotype-specific comparisons. Functional annotation implicated axon guidance, oxidative phosphorylation, and developing midbrain cells. ADHD-to-BPD MR was positive (22 instruments; inverse-variance weighted beta = 0.487, SE = 0.082, p = 3.27 × 10^-9), persisting after outlier removal, MR-LAP correction, and exclusion of two instruments at colocalized loci. Reverse MR was positive (five instruments; beta = 0.303, SE = 0.112, p = 6.83 × 10^-3) but heterogeneous and variant-sensitive. CAUSE favored directional over sharing-only models in both directions (delta ELPD = 6.98 and 5.92, respectively). SuSiE colocalization supported shared signals at rs17576773, rs9969232, and rs4727799.

**Conclusions:** ADHD and BPD exhibit extensive genetic overlap involving 48 shared loci and convergent evidence of directional associations. Findings across complementary methods more consistently support an ADHD-to-BPD relationship, suggesting that shared genetic susceptibility alone may not fully explain their co-occurrence.

## INTRODUCTION

Attention-deficit/hyperactivity disorder (ADHD) and borderline personality disorder (BPD) are distinct psychiatric disorders that frequently co-occur, causing substantial functional and psychosocial impairments. ADHD and BPD affect approximately 1.6–5% and 2.4% of the general population, respectively (Popit et al., 2024; Ramos-Suárez et al., 2026). ADHD is a neurodevelopmental disorder characterized by persistent inattention and/or hyperactivity– impulsivity (Ditrich et al., 2021). Conversely, BPD is characterized by pervasive instability in relationships, self-image, and affect, alongside marked impulsivity (Ditrich et al., 2021). Despite distinct classifications, they share clinically relevant features, particularly impulsivity and emotion dysregulation, complicating differential diagnosis in adulthood (Weiner et al., 2019).

Beyond phenotypic overlap, ADHD and BPD may share vulnerability factors. Genetic predisposition and environmental exposure, such as childhood adversity, have been implicated in both disorders (Distel et al., 2011). Longitudinal studies have shown a higher occurrence of BPD among individuals with childhood ADHD, suggesting that persistent impulsivity and emotion dysregulation contribute to later BPD vulnerability (Tiger et al., 2022). However, clinical overlap makes it difficult to determine whether this association reflects directional developmental pathways or shared liability. Observational studies cannot fully distinguish these possibilities due to potential confounding by unmeasured familial, environmental, and developmental factors.

Genetic methodologies offer a framework for examining the relationship between these disorders. Both conditions exhibit heritability, and recent genome-wide association studies (GWAS) demonstrated significant genetic overlap between ADHD and BPD, indicating that their co-occurrence may be partially attributed to shared genetic susceptibility (Streit et al., 2026). Nevertheless, genetic correlation alone cannot differentiate between shared polygenic architecture, locus-specific overlap, or directional relationships. Conjunctional false discovery rate (conjFDR) analysis identifies loci that are jointly associated by utilizing polygenic overlap (Smeland et al., 2020). In contrast, Mendelian randomization (MR) assesses directional associations, while colocalization analysis determines whether regional associations share a common genetic signal (Burgess et al., 2023). Accordingly, this study aimed to characterize the genetic relationship between ADHD and BPD by examining shared loci and evaluating whether liability to either disorder is directionally associated with the risk of the other. We applied conjFDR to identify jointly associated loci, then conducted bidirectional MR using GWAS summary statistics with sensitivity analyses and Steiger testing. Finally, we integrated locus-specific colocalization and SuSiE fine-mapping to assess shared genetic signals and tested whether MR associations persisted after excluding instruments within these regions.

## METHODS

### Data source and Selection of genetic instruments

This study adhered to the recommendations for MR investigations as outlined by Burgess et al. (2023) and was based exclusively on publicly-available, de-identified datasets. Ethical approval and informed consent were obtained from the original studies in accordance with their respective institutional requirements. We used summary statistics from the largest GWAS meta-analyses of ADHD and a recent BPD GWAS meta-analysis, both available through the Psychiatric Genomics Consortium. The ADHD statistics provided by Demontis et al. (2023) included 38,691 cases and 186,843 controls of European descent (Supplementary Table 1). The BPD statistics, sourced from Streit et al. (2026), comprised 12,339 cases and 1,041,717 controls of European ancestry (Supplementary Table 2).

### Statistical Analysis

We applied linkage disequilibrium score regression (LDSC) to estimate the genome-wide genetic correlation between ADHD and BPD. SNP heritability estimates were converted to liability scale assuming population prevalences of 5% for ADHD and 1.5% for BPD, accounting for effective sample sizes as a balanced case-control sample (effective sample prevalence = 0.5). To assess polygenic enrichment between phenotype pairs, we examined conditional quantile-quantile plots of the p-value distribution for one phenotype given p-value thresholds for another (p < 0.1, p < 0.01, and p < 0.001). To limit inflation, the major histocompatibility complex region was excluded. Shared loci were identified using conjFDR analysis, considering SNPs with conjFDR < 0.05 as shared (Smeland et al., 2020). To evaluate shared-loci consistency, we performed sign-concordance analyses (Karadag et al., 2023) using FinnGen Release 13 (R13) F5_ADHD and F5_EMOPER statistics (Kurki et al., 2023).

Inspection of the BPD discovery GWAS cohort revealed sample overlap from inclusion of FinnGen Release 8 (R8) F5_EMOPER data. To address this, and because no sufficiently independent BPD GWAS datasets were available, we used MetaSubtract (Nolte et al., 2020) to remove the FinnGen R8 F5_EMOPER contribution from the Streit et al. meta-analysis, generating an R8-depleted BPD GWAS for directional validation.

Sign concordance was assessed by comparing (i) ADHD effect estimates from the Demontis et al. discovery GWAS with FinnGen R13 F5_ADHD and (ii) BPD effect estimates from the FinnGen R8-depleted Streit et al. GWAS with FinnGen R13 F5_EMOPER. After harmonization, we calculated the proportion of lead variants with concordant beta signs. Significance was assessed using two-sided binomial tests. Variants that were unavailable, could not be reliably harmonized, or showed discordant effects in either comparison were not considered jointly replicated.

To elucidate the biological mechanisms contributing to the shared genetic architecture of ADHD and BPD, conjFDR-loci were functionally characterized and mapped to putative genes using Functional Mapping and Annotation of Genome-Wide Association Studies (FUMA; Watanabe et al., 2017). These genes were then examined through functional enrichment analyses of Gene Ontology terms and Kyoto Encyclopedia of Genes and Genomes pathways, as well as analyses of tissue- and cell type-specific expression.

Directional relationships between the two traits were explored using a MR framework. In MR statistics, genetic instruments were selected from the corresponding GWAS at genome-wide significance (p < 5 × 10^-8). Variants were clumped using the European subset of the 1000 Genomes Project Phase 3 as the LD reference, with r^2 < 0.001 within a 10-Mb window. Only variants with adequate strength (F > 10) were retained.

Primary MR analysis used inverse-variance weighted multiplicative random-effects models (IVW) to account for between-instrument heterogeneity, confirmed in fixed-effects analyses (Cochran’s Q p < 0.05 in both directions). We also used the weighted-median estimator, which can provide consistent estimates if up to 50% of the instrument weight comes from invalid instruments (Bowden et al., 2016). Heterogeneity was assessed using Cochran’s Q and I^2 statistics (Burgess et al., 2023). We performed leave-one-out analyses (LOO), MR-Egger, MR pleiotropy residual sum and outlier (MR-PRESSO; Verbanck et al., 2018), and radial-MR. Steiger directionality testing across multiple plausible prevalence scenarios for ADHD and BPD assessed whether instruments explained more phenotypic variance in the designated exposure than outcome (Burgess et al., 2023). MR-LAP addressed potential bias from sample overlap, weak instruments, and winner’s curse (Mounier & Kutalik, 2023). Furthermore, Causal Analysis Using Summary Effect estimates (CAUSE; Morrison et al., 2020) was performed to distinguish causal effects from correlated horizontal pleiotropy.

We also investigated whether MR findings reflected local genetic architecture using Bayesian colocalization with the coloc framework (Giambartolomei et al., 2014). All instruments were tested bidirectionally across genomic regions (+/-500 kb) centered on lead SNPs using effect estimates, variances, allele frequencies, and sample-size information from GWAS summary statistics.

To assess the statistical sensitivity of the bidirectional MR analyses and contextualize differences in instrument strength and availability between the two directions, we calculated the minimum detectable effects (MDEs) corresponding to 80% power at a two-sided significance threshold of alpha = 0.05. Using SNP-level associations, fixed-effects inverse-variance weighted (IVW) standard errors were calculated and adjusted for heterogeneity under a multiplicative random-effects model using max(1, Q/[k-1]), where Q is Cochran’s heterogeneity statistic and k the number of instruments. Log-odds MDEs used (z_0.975 + z_0.80) x SE, were exponentiated and interpreted as conditional detection thresholds.

Genome-wide significance was defined as p < 5 × 10^-8 for instrument selection. For MR analyses, statistical significance was defined as a two-sided p < 0.05. The MR-PRESSO global test and Steiger directionality test were evaluated at p < 0.05. For colocalization analyses, posterior probabilities rather than p-value thresholds were used, with PP.H4 > 0.80 considered strong evidence of colocalization. A detailed description of the statistical pipeline and software packages used is provided in the supplementary methods section.

## RESULTS

### Genome-wide genetic correlation between ADHD and BPD

LDSC revealed a genetic correlation between ADHD and BPD (r_g = 0.733, SE = 0.031, P = 6.73 × 10^-122), indicating shared variant influences. SNP-based heritability estimates using sample sizes were 0.165 (SE = 0.008) for ADHD and 0.281 (SE = 0.018) for BPD; on the liability scale, estimates were 0.140 (SE = 0.006) and 0.171 (SE = 0.011), assuming prevalences of 5% and 1.5%, respectively. LDSC intercepts were 1.036 (SE = 0.010) for ADHD and 1.036 (SE = 0.009) for BPD; the cross-trait genetic covariance intercept was 0.026 (SE = 0.007). The BPD heritability estimate matched Streit et al. (liability-scale h^2 = 0.173), supporting comparability with the GWAS.

### Cross-trait genetic enrichment

Conditional Q-Q plots revealed bidirectional cross-trait enrichment between ADHD and BPD. In both directions, more stringent thresholds for the conditioning phenotype corresponded to leftward deviations from the null expectation, indicating shared associations (Supplementary Figure S1 panels A and C). Conditional fold-enrichment analyses showed increasing enrichment across conditioning thresholds, corroborating enrichment (Supplementary Figure S1 panels B and D). ConjFDR analysis evidenced 48 shared loci (Figure 1). To assess consistency, we evaluated 48 SNPs using estimates from the ADHD discovery GWAS, the BPD GWAS following the MetaSubtract procedure, and FinnGen R13 F5_ADHD. Of the 48 loci, 37 (77.1%) showed concordant directions between the ADHD discovery GWAS and FinnGen R13 F5_ADHD (two-sided exact binomial p = 2.22 × 10^-4); 40 of 48 loci (83.3%) showed concordant directions between the FinnGen R8-depleted Streit BPD GWAS and FinnGen R13 F5_EMOPER (two-sided exact binomial p = 3.31 × 10^-6). Concordance in both phenotype-specific comparisons was observed for 36 of the 48 lead variants (75.0%), representing the intersection of the two concordant sets.

**Figure 1.**
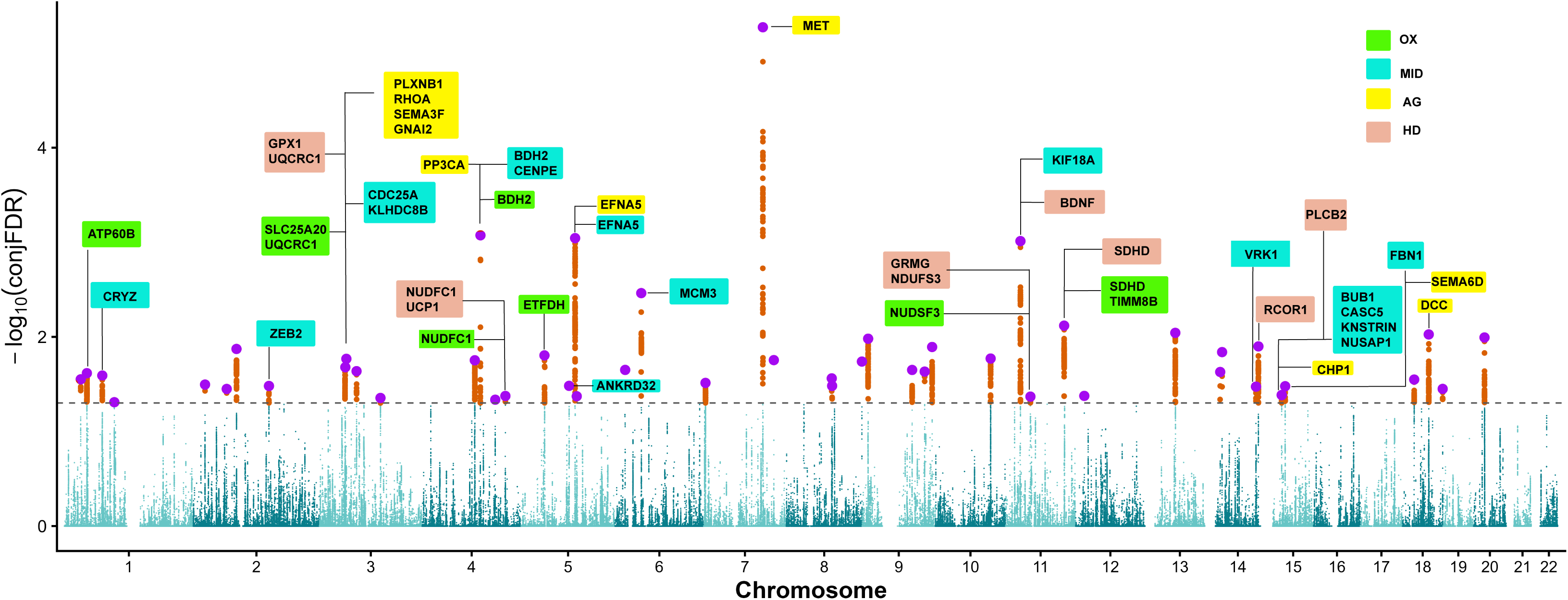
Conjunctional false discovery rate analysis of ADHD and BPD. Manhattan plot showing -log (conjFDR) values across the 22 autosomes. The dashed horizontal line indicates the significance threshold of conjFDR < 0.05. Orange points represent variants exceeding this threshold, and purple points highlight selected lead variants. Labels indicate selected genes mapped by FUMA to shared loci that overlapped with gene sets identified in downstream enrichment analysis. AG, axon guidance; MD, Manno_Midbrain_Neurotypes_HPROGFPL; OX, oxidative phosphorylation; HD, Huntington’s disease

**Table 1.** All distinct ADHD loci associated with BPD at conjFDR < 0.05. P-values are reported for the ADHD phenotype. A1 is the ADHD effect allele. Signs of Z-values for both traits are oriented to the ADHD A1 allele. The Validation column indicates joint directional concordance of genetic effects for ADHD and BPD in the FinnGen-based validation analysis (Yes = concordant; No = non-concordant). Overall, 36 of 48 loci (75%) showed joint directional concordance.

| CHR | Lead SNP | Position | Nearest gene | Distance (bp) | A1/A2 | p-value | conjFDR | Validation |
|---|---|---|---|---|---|---|---|---|
| 1 | rs3766823 | 32197257 | BAI2 | 0 | G/A | 4.98e-05 | 0.0281 | Yes |
| 1 | rs3791151 | 44078602 | PTPRF | 0 | C/A | 6.65e-09 | 0.0242 | Yes |
| 1 | rs35641559 | 73760104 | RP4-598G3.1 | 11748 | T/C | 7.74e-05 | 0.0256 | Yes |
| 1 | rs6593608 | 96987688 | RN7SL831P | 61076 | C/T | 8.88e-05 | 0.049 | Yes |
| 2 | rs12620364 | 22562609 | AC068490.2 | 0 | G/A | 0.000117 | 0.0319 | Yes |
| 2 | rs10496105 | 64320746 | PELI1 | 0 | G/A | 0.000143 | 0.0355 | Yes |
| 2 | rs11904603 | 83165014 | AC098817.5 | 80128 | T/C | 2.3e-05 | 0.0134 | No |
| 2 | rs726857 | 145753424 | TEX41 | 0 | T/A | 8.37e-07 | 0.033 | No |
| 3 | rs6446298 | 49860854 | AC139451.1 | 4367 | T/C | 5.2e-05 | 0.0209 | No |
| 3 | rs7634751 | 51846733 | RP11-314A5.3 | 4885 | G/C | 1.7e-06 | 0.0171 | Yes |
| 3 | rs56207020 | 71486387 | FOXP1 | 0 | G/A | 6.3e-05 | 0.023 | No |
| 3 | rs113786703 | 117602013 | LSAMP:RP11-384F7.2 | 0:0 | G/T | 8.16e-07 | 0.0442 | Yes |
| 4 | rs2173844 | 101489327 | EMCN | 0 | C/T | 1.83e-05 | 0.0177 | Yes |
| 4 | rs72678864 | 112422145 | RP11-255I10.1 | 50864 | G/A | 5.35e-10 | 0.000844 | Yes |
| 4 | rs55669488 | 140943275 | MAML3 | 0 | T/A | 0.000152 | 0.0461 | No |
| 4 | rs9995801 | 160489178 | U3 | 60997 | G/A | 8e-05 | 0.0421 | Yes |
| 5 | rs113901452 | 44816452 | MRPS30 | 0 | G/A | 4.43e-07 | 0.0157 | Yes |
| 5 | rs34523 | 92303352 | CTC-458G6.2 | 28053 | T/C | 3.84e-07 | 0.0329 | Yes |
| 5 | rs2431112 | 103931707 | RP11-6N13.1 | 0 | G/A | 2.04e-10 | 0.000904 | Yes |
| 5 | rs6882236 | 107092298 | RN7SL782P | 21530 | T/C | 0.000102 | 0.0424 | Yes |
| 6 | rs6940685 | 19700166 | RP1-167F1.2:RP3-471C18.2 | 0:0 | T/C | 4.35e-05 | 0.0223 | Yes |
| 6 | rs78648104 | 50683009 | TFAP2D | 0 | T/C | 1.24e-07 | 0.00345 | Yes |
| 7 | rs73293551 | 3550676 | SDK1 | 0 | A/T | 0.000108 | 0.0306 | No |
| 7 | rs4727799 | 114110568 | FOXP2 | 0 | C/T | 8.91e-11 | 5.33e-06 | Yes |
| 7 | rs10258495 | 135248558 | NUP205 | 0 | T/C | 3.81e-05 | 0.0176 | Yes |
| 8 | rs72696111 | 87773943 | RP11-386D6.1 | 0 | A/G | 3.04e-05 | 0.0274 | Yes |
| 8 | rs59406400 | 88462804 | CNBD1 | 0 | A/G | 6.58e-06 | 0.033 | Yes |
| 8 | rs4925813 | 145856021 | ARHGAP39 | 0 | A/G | 2.51e-07 | 0.0182 | Yes |
| 9 | rs4262394 | 11599813 | RP11-23D5.1 | 323498 | A/G | 1.46e-05 | 0.0105 | Yes |
| 9 | rs7857266 | 96381765 | PHF2 | 0 | C/T | 5.97e-05 | 0.0224 | Yes |
| 9 | rs10739498 | 120513796 | snoZ13_snr52 | 20046 | G/A | 6.42e-05 | 0.0233 | Yes |
| 9 | rs7026534 | 134907263 | MED27 | 0 | T/G | 1.11e-05 | 0.0128 | No |
| 10 | rs2491381 | 106620758 | SORCS3 | 0 | A/C | 1.29e-05 | 0.0169 | Yes |
| 11 | rs10767735 | 28642561 | RP11-960D24.1 | 58168 | G/C | 2.27e-12 | 0.000971 | No |
| 11 | rs10466480 | 47889737 | NUP160 | 19629 | C/T | 3.23e-06 | 0.0428 | Yes |
| 11 | rs2298527 | 112851961 | NCAM1 | 0 | G/C | 8.2e-06 | 0.00761 | No |
| 12 | rs10846378 | 16743434 | MGST1:LMO3 | 0:0 | T/G | 5.76e-05 | 0.0421 | No |
| 13 | rs1334297 | 58335375 | PCDH17 | 31929 | G/A | 1.11e-05 | 0.00906 | Yes |
| 14 | rs12894091 | 29752200 | RP11-562L8.1 | 0 | G/A | 4.32e-05 | 0.0235 | Yes |
| 14 | rs35985675 | 33391029 | NPAS3 | 13109 | T/A | 4.88e-06 | 0.0145 | No |
| 14 | rs77653640 | 98643863 | RP11-61O1.1 | 0 | G/A | 8.56e-09 | 0.0336 | Yes |
| 14 | rs11629241 | 103379225 | TRAF3 | 1387 | T/C | 8.06e-06 | 0.0126 | Yes |
| 15 | rs12912058 | 40903814 | CASC5 | 0 | C/A | 7.28e-05 | 0.0412 | No |
| 15 | rs281265 | 47675602 | SEMA6D:CTD-2050N2.1 | 0:0 | C/A | 2.75e-05 | 0.0332 | Yes |
| 18 | rs4132769 | 22695332 | ZNF521 | 0 | A/G | 9.25e-05 | 0.0282 | Yes |
| 18 | rs7245004 | 50716945 | DCC | 0 | C/A | 5.4e-08 | 0.00942 | Yes |
| 18 | rs71367544 | 77574374 | RP11-154H12.3 | 0 | C/T | 8.2e-06 | 0.0355 | Yes |
| 20 | rs6047233 | 21059248 | LINC00237 | 8996 | C/A | 6.57e-09 | 0.0101 | Yes |

### Functional Annotation

Functional mapping of the 48 shared ADHD-BPD loci (Supplementary Data Sheets 1-7) identified 629 mapped genes, corresponding to 430 unique Entrez genes after deduplication. 131 candidate SNPs had CADD scores exceeding 12.37, indicating predicted deleteriousness. Among 198 significantly enriched gene sets (adjusted p < 5.0 × 10^-2), four KEGG/Hallmark pathways were identified. Kyoto Encyclopedia of Genes and Genomes (KEGG) Axon guidance showed the strongest statistical evidence of enrichment among these pathways, with 10 overlapping genes among 127 pathway genes (4.42-fold enrichment; p = 9.47 × 10^-5; adjusted p = 1.76 × 10^-2). KEGG Glycosaminoglycan degradation included 4 of 19 genes and showed the largest relative enrichment (11.82-fold; p = 3.11 × 10^-4; adjusted p = 2.89 × 10^-2), although based on a four-gene overlap. KEGG Huntington’s disease included 10 of 164 genes, and Hallmark Oxidative Phosphorylation included 11 of 195 genes. Genes contributing to Axon guidance and Huntington’s disease enrichments were distributed across seven loci, whereas Glycosaminoglycan degradation mapped to a single region, indicating that this signal was locus-driven rather than supported by convergence across independent loci. Hallmark Oxidative Phosphorylation genes were distributed across eight loci, with two mapping to locus 9 (Figure 1). Cell-type signature analysis identified 2.91-fold enrichment for the MANNO_MIDBRAIN_NEUROTYPES_HPROGFPL signature (16 of 309 genes; adjusted p = 2.48 × 10^-2), with overlapping genes distributed across 11 loci (Figure 1).

### Bidirectional Mendelian Randomization

Using the full BPD GWAS dataset, 1,428 ADHD variants reached genome-wide significance (p < 5 × 10^-8). After LD clumping, 26 independent variants were retained as strong candidate genetic instruments (F-statistics: 29.85-60.13; mean, 38.89; median, 34.85; Supplementary table 5). Of the 26 instruments, 22 were directly available and could be harmonized with the BPD GWAS (Supplementary table 6). Multiplicative random-effects IVW analysis showed a positive association between genetic liability to ADHD and BPD (figure 2, panel A; p = 3.3 × 10^-9). Heterogeneity existed among SNP-specific estimates (Q = 51.09; df = 21; p = 2.57 × 10^-4; I^2 = 58.9%), but 17 of 22 Wald ratio estimates were directionally concordant (Supplementary Figure 2). MR-Egger regression yielded a positive but imprecise slope (Figure 2, panel A; p = 0.151). The MR-Egger intercept did not provide statistically significant evidence of directional horizontal pleiotropy (intercept = −0.0009, SE = 0.0219, p = 9.67 × 10^-1; Supplementary Figure 3). LOO indicated that the ADHD-to-BPD association was not driven by any single genetic instrument (Supplementary Figure 5).

**Figure 2.**
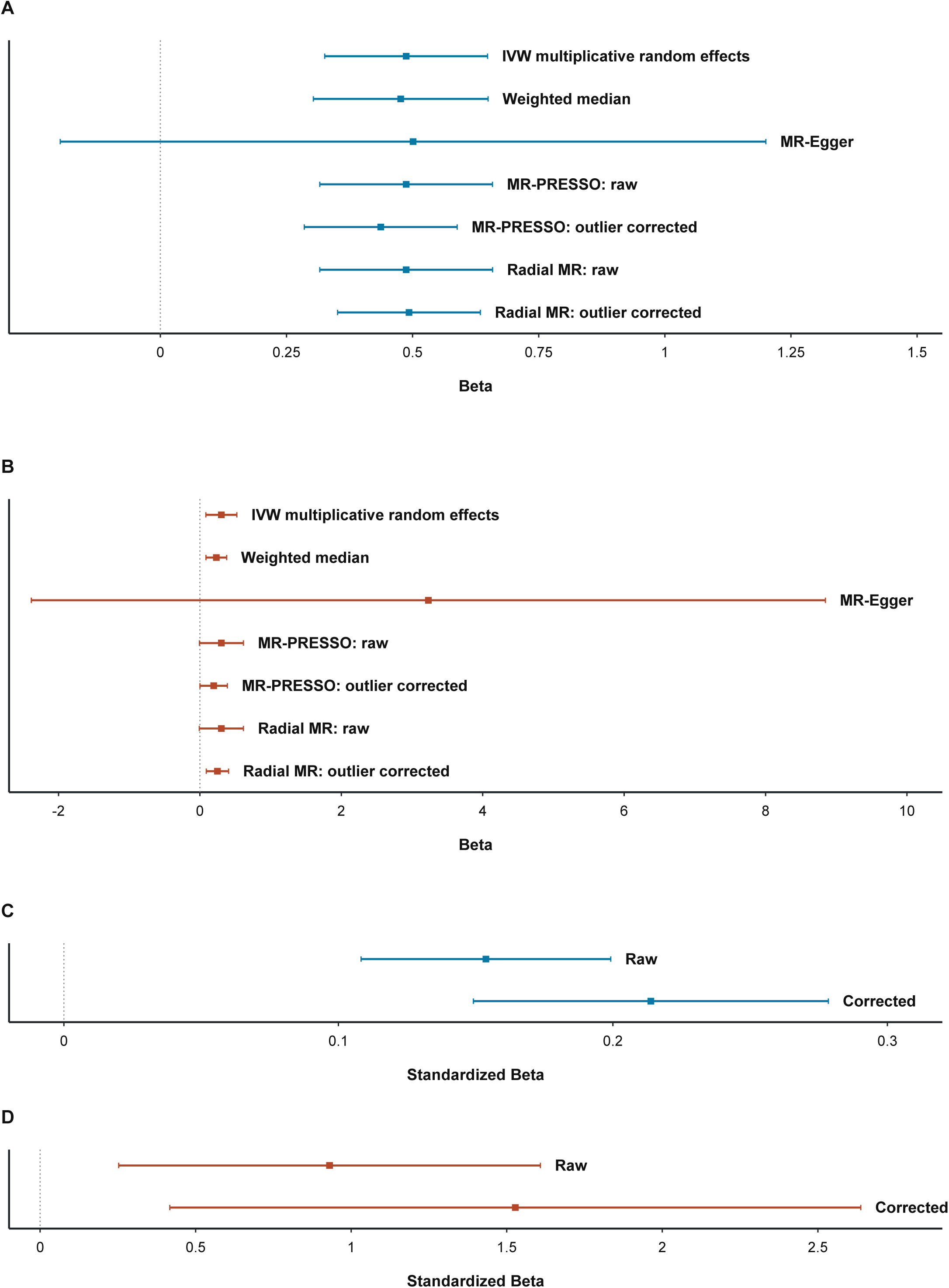
Bidirectional Mendelian randomization analyses of ADHD and BPD. Forest plots show effect estimates for ADHD → BPD (A) and BPD → ADHD (B), obtained using multiplicative random-effects inverse-variance weighting (IVW), weighted median, MR-Egger, MR-PRESSO, and Radial MR. Raw and outlier-corrected estimates are shown for MR-PRESSO and Radial MR. Panels C and D display observed and bias-corrected MR-LAP estimates in both directions.

Radial MR (Supplementary Figure 6) yielded consistent estimates (t = 5.916; p = 7.15 × 10^-6; Figure 2, panel A). After excluding three outliers (rs9969232, rs2886697, and rs10875612), the association remained significant across 19 instruments (t = 7.320; p = 8.48 × 10^-7; Figure 2, panel A) and residual heterogeneity became non-significant (Q = 22.73, df = 18, p = 2.01 × 10^-1, I^2 = 20.81%). Consistent with the Radial MR findings, MR-PRESSO detected global pleiotropy (Global Test RSSobs = 56.32; p = 1.4 × 10^-3) and identified one outlier (rs9969232). After outlier correction, the association remained significant (t = 6.008; p = 7.11 × 10^-6; Figure 2, panel A), with no significant difference between raw and corrected estimates (distortion test p = 3.94 × 10^-1). Reversely, five of the six identified BPD instruments were analyzed (Supplementary Tables 7 and 8). Genetic liability to BPD (Figure 2, panel B) was positively associated with ADHD using multiplicative random-effects IVW (Z = 2.705; p = 6.83 × 10^-3) and weighted median estimators (Z = 3.101; p = 1.93 × 10^-3).

Substantial between-instrument heterogeneity was observed (Q = 20.24, df = 4, p = 4.47 × 10^-4; I^2 = 80.2%). The MR-Egger intercept did not differ significantly from zero (intercept = −0.251, SE = 0.151; t = −1.662, df = 3; p = 1.95 × 10^-1; Supplementary Figure 8). However, the MR-Egger slope was highly imprecise (t = 1.831, df = 3; p = 1.64 × 10^-1; Figure 2, panel B), and its interpretation was limited by the small number of instruments. However, also in BPD->ADHD direction, LOO indicated that the association was not driven by any single genetic instrument (Supplementary figure 10).

MR-PRESSO identified rs4727799 as an outlier (global test p = 1 × 10^-2); its exclusion attenuated the association, which remained positive (Figure 2, panel B; p = 4.95 × 10^-2) with significant distortion compared with the uncorrected estimate (p < 1 × 10^-4). Radial MR using modified second-order weights showed a non-significant positive association between genetic liability to BPD and ADHD across five instruments (t = 2.704, df = 4; p = 5.39 × 10^-2; Figure 2, panel B), with significant heterogeneity (Q = 16.68, df = 4, p = 2.23 × 10^-3; I^2 = 76.0%). After excluding two outliers (rs4727799, rs6549383), the association was significant across three retained instruments (t = 6.735, df = 2; p = 2.13 × 10^-2; Supplementary Figure 11), with non-significant residual heterogeneity (Q = 0.57, df = 2, p = 7.54 × 10^-1; I^2 = 0%). These findings support a reverse-direction association, though limited by few instruments and post-hoc outlier exclusion.

To evaluate sample overlap and winner’s curse, we applied MR-LAP. For ADHD->BPD (26 instruments; Supplementary Table 9), the observed IVW showed a positive association (Figure 2 panel C, p = 3.50 × 10^-11). After bias correction, the association remained significant (Figure 2 panel D, p = 8.37 × 10^-11), with a significant difference between estimates (p = 1.36 × 10^-9). The MR-Egger intercept showed no directional pleiotropy (p = 0.133). For BPD->ADHD (5 instruments; Supplementary Table 10), both observed (p = 7.0 × 10^-3) and corrected estimates (p = 8.0 × 10^-3) were significant. The MR-Egger intercept was not significant (p = 3.7 × 10^-1), although its interpretability was limited by the small number of instruments. Comparison with conventional MR instruments demonstrated substantial concordance in both directions.

Across 20 combinations of assumed BPD and ADHD population prevalences, Steiger analyses continued to support the intended orientation of the instruments in both directions (Supplementary Tables 11 and 12).

### CAUSE analysis

To distinguish directional causal effects from correlated horizontal pleiotropy, we applied CAUSE. After harmonizing ADHD and BPD GWAS datasets, 4,729,831 variants were available. Exposure-associated variants (P < 1 × 10^-3) were LD-pruned at r^2 < 0.1 using the CAUSE LD reference panel, yielding 2,135 variants for ADHD-to-BPD and 1,619 for BPD-to-ADHD. The correlation between estimation errors was rho = 0.181 and was incorporated into both models.

In the ADHD-to-BPD direction, both the sharing model (delta ELPD [sharing - null] = 52.49, SE = 8.60) and the causal model (delta ELPD [causal - null] = 59.47, SE = 9.74) outperformed the null model. The causal model fit better than the sharing model (delta ELPD [causal - sharing] = 6.98, SE = 1.33, P = 7.3 × 10^-8), with posterior median gamma = 0.27 (95% CrI, 0.21-0.32).

In the BPD-to-ADHD direction, both models outperformed the null model (sharing: delta ELPD = 29.46, SE = 6.53; causal: delta ELPD = 35.37, SE = 7.82). The causal model again fit better than the sharing model (delta ELPD [causal - sharing] = 5.92, SE = 1.54, P = 6.2 × 10^-5), with posterior median gamma = 0.16 (95% CrI, 0.12-0.20).

Sharing-only models estimated eta = 0.35 (95% CrI, 0.28-0.45) and q = 0.65 (95% CrI, 0.48-0.81) for ADHD-to-BPD, and eta = 0.26 (95% CrI, 0.18-0.37) and q = 0.51 (95% CrI, 0.31-0.71) for BPD-to-ADHD. Although the sharing-only models indicated substantial correlated pleiotropy, the causal model provided better predictive performance in both directions, suggesting that directional effects accounted for variation beyond that explained by correlated horizontal pleiotropy alone.

### Colocalization analysis

In the ADHD->BPD direction, coloc-ABF showed strongest evidence of a shared association signal at rs9969232 (PP.H4 = 0.990), rs17576773 (PP.H4 = 0.972) and rs2582895 (PP.H4 = 0.875), with additional support at rs11596214 (PP.H4 = 0.616) and rs7506904 (PP.H4 = 0.515). In contrast, rs17718444 and rs76284431 supported distinct signals (Supplementary Table 13). SuSiE-based fine-mapping identified 25 credible sets across 19 loci: 21 for ADHD and four for BPD. Fifteen loci yielded ADHD-only credible sets, whereas four (rs17718444, rs17576773, rs9969232, and rs76284431) yielded sets for both traits (Supplementary Table 14). Two ADHD loci, rs7844069 and rs2582895, contained two credible sets each; posterior coverage ranged from 95.2% to 99.5%. Among the four loci evaluable by coloc-SuSiE, two confirmed shared signals. The chromosome 4 locus indexed by rs17576773 yielded PP.H4 = 0.972, with 17 ADHD-associated and four BPD-associated variants, including three shared variants. The chromosome 7 locus indexed by rs9969232 showed PP.H4 = 0.989, with 46 and 37 variants in the ADHD and BPD credible sets, including 30 shared variants. At chromosome 4, rs72678864 had the highest BPD posterior inclusion probability among shared variants (PIP = 0.486; ADHD PIP = 0.050). At chromosome 7, rs9969232 showed PIPs of 0.372 for ADHD and 0.165 for BPD, whereas rs4727799 showed 0.048 and 0.466; rs4727799 was an instrument in the reverse BPD-to-ADHD MR analysis. In contrast, coloc-SuSiE favored distinct signals at rs17718444 (PP.H3 = 0.989; PP.H4 = 0.0007) and rs76284431 (PP.H3 = 0.976; PP.H4 = 0.0009). The remaining 18 regions could not be evaluated because sets were not identified for both traits (Supplementary Table 14). In the BPD->ADHD analysis, coloc-ABF across five loci showed colocalization at rs4727799 (PP.H4 = 0.990), distinct signals at rs6549383 (PP.H3 = 0.982), and H2 support at three loci (Supplementary Table 15). Coloc-SuSiE confirmed colocalization at rs4727799 (PP.H4 = 0.985), whereas the signals at rs6549383 were distinct (PP.H3 = 0.989). Similar to the ADHD-to-BPD direction, the remaining regions could not be formally evaluated using coloc-SuSiE due to the absence of identified credible sets for both traits (Supplementary Table 16).

### Sensitivity analyses excluding lead SNPs from colocalized loci

To assess whether bidirectional MR estimates were influenced by variants at loci colocalized in the SuSiE analyses, we repeated the analyses after excluding corresponding lead SNPs. Excluding rs9969232 and rs17576773 did not materially alter the ADHD->BPD association. The IVW and weighted median estimates remained positive and statistically significant. Additional radial-MR analyses identified nominal outliers, but exclusion of these variants also retained a positive association with reduced heterogeneity (Supplementary tables 17-18). In the BPD->ADHD direction, exclusion of rs4727799 provided estimations consistent with the primary analysis, with no evidence of heterogeneity. However, these results were cautiously interpreted, given the limited number of instruments.

### Minimum Detectable effect analysis

At 80% power and a two-sided significance threshold of alpha = 0.05, the minimum detectable odds ratio (MDE), calculated using multiplicative random-effects inverse-variance weighted (IVW) standard errors, was 1.259 for ADHD->BPD with 22 genetic instruments, compared with an observed IVW odds ratio (OR) of 1.628. Following sequential instrument exclusions, the MDE decreased to 1.230 with 20 instruments and 1.220 with 18 instruments, while the corresponding observed ORs were 1.518 and 1.509, respectively. In the reverse direction (BPD->ADHD), the conditional MDE was 1.369 with five instruments, compared with an observed IVW OR of 1.354, in the presence of substantial between-instrument heterogeneity (Cochran’s Q = 20.24, df = 4, p = 4.47 × 10^-4). After excluding rs4727799, the MDE decreased to 1.186 with four instruments, while the observed IVW OR was 1.215 and residual heterogeneity was no longer statistically significant (Q = 3.48, df = 3, p = 3.23 × 10^-1).

## DISCUSSION

This study combined conjFDR and bidirectional MR to examine the shared polygenic architecture and possible directional genetic associations between ADHD and BPD. We identified 48 loci associated with both disorders. Analyses suggested a positive association between genetic liability to ADHD and BPD, while evidence for the reverse direction was less precise and more sensitive to individual variants.

The primary multiplicative random-effects IVW analysis supported an association between ADHD and BPD, with directionally consistent results across sensitivity analyses. Although the instruments showed heterogeneity, the MR-Egger intercept provided no evidence of directional horizontal pleiotropy, and LOO indicated that no single instrument drove the estimate. The association remained positive after excluding radial-MR outliers and eliminating heterogeneity. MR-PRESSO detected pleiotropy but yielded a positive outlier-corrected estimate. MR-LAP also supported a positive ADHD->BPD association after adjusting for sample overlap, winner’s curse, and weak-instrument bias. The corrected estimate remained statistically significant and exceeded the observed standardized IVW estimate, suggesting these biases do not fully explain the association. Steiger analyses also supported the expected direction, with instruments explaining more variance in ADHD than in BPD. However, we cannot completely exclude correlated pleiotropy: ADHD-related variants might influence BPD through related factors such as impulsivity, emotion dysregulation, or broader mental health issues rather than directly through ADHD. Violations of instrument independence due to population differences, selective mating, or genetic confounding are also possible, and the Steiger-test results do not definitively establish causal direction.

Colocalization analyses identified two ADHD instrument loci, rs9969232 and rs17576773, with evidence of shared ADHD->BPD association signals in both coloc-ABF and coloc-SuSiE analyses. After excluding these variants, the IVW association remained positive, suggesting the estimate was not entirely attributable to these two SNPs.

The reverse BPD->ADHD analysis requires cautious interpretation. Although IVW and weighted median analyses indicated a positive association, only five instruments were available with substantial heterogeneity. The association was influenced by rs4727799, a chromosome 7 variant showing colocalization between ADHD and BPD; removing it reduced association magnitude and heterogeneity, although the estimate remained positive. Similarly, radial-MR analyses identified rs4727799 and rs6549383 as outliers; after exclusion, the association remained positive but relied on only three instruments.

The reverse-direction result is less stable than the ADHD->BPD result and may reflect the shared genetic architecture at the chromosome 7 locus together with limited instrument precision. Given limited instrument number and sensitivity to individual variants, the reverse-direction findings do not establish a direct developmental pathway from BPD to ADHD and may partly reflect shared genetic liability underlying dimensional traits such as impulsivity and emotional dysregulation.

CAUSE analyses offered a complementary genome-wide assessment of the genetic overlap between ADHD and BPD. In both directions the causal model fit the data significantly better than the sharing-only model, suggesting correlated horizontal pleiotropy alone did not fully account for the observed association patterns under CAUSE assumptions. The estimated directional effect was more pronounced from ADHD to BPD than from BPD to ADHD; however, differences in GWAS characteristics prevent direct comparisons of effect sizes. Additionally, directional-model support in the BPD-to-ADHD analysis contrasted with the greater sensitivity to individual instruments in conventional MR, with only five BPD instruments. This discrepancy likely arises from instrument selection: CAUSE employs genome-wide variants (p < 1 × 10^-3), whereas MR uses genome-wide significant instruments (p < 5 × 10^-8). The CAUSE findings suggest directional effects at the genome-wide level, but the limited number of BPD instruments restricts the precision of conventional MR in the reverse direction.

The conjFDR-identified shared loci provide a further perspective on the ADHD-BPD relationship. Functional annotation implicated pathways related to axon guidance, oxidative phosphorylation, and neurodevelopmentally relevant cell type signatures. The axon-guidance and oxidative phosphorylation findings involved genes distributed across multiple loci, which is more consistent with broader biological convergence than enrichment driven by a single region. In contrast, glycosaminoglycan degradation enrichment was attributable to genes from one locus and should be interpreted as locus-specific rather than as evidence of a general pathway-level mechanism. These annotations are hypothesis-generating and do not establish the causal genes, affected tissues, or molecular mechanisms underlying shared associations. The chromosome 7 region containing FOXP2 represents a particularly notable shared locus. Colocalization and fine-mapping analyses supported a shared association signal in this region, although the posterior inclusion probabilities differed between ADHD and BPD across individual variants. FOXP2 has established roles in neurodevelopmental and corticostriatal processes (Vargha-Khadem et al., 2005), making it a biologically plausible candidate in the context of ADHD and BPD-related phenotypes. Nevertheless, the current analyses do not establish FOXP2 as the causal gene, and the variants identified may affect other genes or regulatory elements within the region. Similarly, chromatin-interaction mapping has implicated MET; however, the interaction evidence available was obtained from mesenchymal stem cells rather than neural tissue. Consequently, any suggested role of MET in the ADHD-BPD association should be considered provisional, despite its previous implication in neurodevelopmental processes (Campbell et al., 2006; Ahirwar et al., 2025).

This study has several limitations. First, MR estimates rely on assumptions that cannot be fully verified, including the absence of horizontal pleiotropy and confounding pathways. In this context, developmental pleiotropy (e.g., shared effects on impulsivity or executive functions) is plausible and may violate the exclusion restriction even after colocalization-based exclusion. Although the sensitivity analyses, CAUSE, MR-LAP, and targeted removal of colocalized loci addressed several potential sources of bias, none could conclusively establish causality. The small but statistically significant cross-trait LDSC intercept (0.026, SE = 0.007) suggests modest correlated estimation error between the discovery GWAS, potentially reflecting sample overlap or other sources of covariance. Although MR-LAP and FinnGen R8 depletion addressed sample-overlap concerns in complementary analyses, residual bias cannot be entirely excluded, warranting caution in causal interpretation without negating the consistency of the observed genetic associations. Second, the BPD GWAS yielded fewer genome-wide significant genetic instruments than the ADHD GWAS, constraining the power and precision of the BPD-to-ADHD Mendelian randomization analysis. Minimum detectable effect analyses indicated that the BPD-to-ADHD direction was only powered to detect relatively large effects (OR > 1.37 with five instruments), raising the possibility a true smaller effect was overlooked. Third, the use of European-ancestry GWAS limits generalizability to other populations. Fourth, the BPD phenotype and the FinnGen emotionally unstable personality disorder phenotype may not be fully interchangeable, despite their conceptual overlap. Finally, the sign-concordance analysis supports directional consistency of the shared loci across datasets but does not constitute independent replication of the full conjFDR or MR analyses.

These findings may have clinical relevance. Shared genetic susceptibility may partly explain why ADHD and BPD frequently co-occur and why difficulties in impulsivity and emotion regulation can complicate their differential diagnosis. The stronger and more consistent ADHD-to-BPD findings are compatible with the hypothesis that genetic liability to ADHD is associated with increased BPD risk. Whether this reflects a developmental pathway in which ADHD-related traits precede BPD features, or shared liability to dimensional traits such as impulsivity, requires validation in longitudinal cohorts with repeated phenotyping. This does not imply that ADHD inevitably leads to BPD or that genetic risk establishes individual-level clinical progression. Rather, our results may reinforce the clinical relevance of monitoring emotional dysregulation, impulsivity, interpersonal difficulties, and emerging borderline personality features during adolescence and early adulthood among individuals with ADHD. Future studies using larger and more diverse BPD GWAS, longitudinal cohorts, and functional genomic data are needed to clarify the biological mechanisms underlying these shared and potentially directional genetic relationships.

## Supporting information

Supplementary Data Sheets 1-7

supplementary methods section.

## DATA AVAILABILITY

The genome-wide association study (GWAS) summary statistics for attention-deficit/hyperactivity disorder (ADHD) and borderline personality disorder (BPD) were obtained from publicly available datasets associated with Demontis et al. (2023; https://doi.org/10.6084/m9.figshare.22564390) and Streit et al. (2026; https://doi.org/10.6084/m9.figshare.30777305), respectively. The FinnGen GWAS summary statistics used in this study, including releases R8 and R13, are publicly available through the FinnGen data access portal (https://www.finngen.fi/en/access_results). Access to FinnGen summary statistics requires completion of an online access form. All analyses were conducted using aggregated GWAS summary statistics; no individual-level participant data were accessed.

## FUNDING

This research did not receive any specific grant from funding agencies in the public, commercial, or not-for-profit sectors.

## DECLARATION OF COMPETING INTERESTS

The authors declare that they have no known competing financial interests or personal relationships that could have appeared to influence the work reported in this paper.

## ACKNOWLEDGEMENTS

We acknowledge the participants and investigators of the FinnGen study and Psychiatric Genomics Consortium.

## Notes

### Competing Interest Statement

The authors have declared no competing interest.

### Summary of Updates

Two minor typographical and editorial corrections were made in the Discussion section The typographical error his study was corrected to This study. The phrase Several limitations apply was replaced with This study has several limitationsfor improved clarity. These changes are purely editorial and do not affect the scientific content, methodology, results, or conclusions of the study.

