## supplementary methods section. for "SHARED GENETIC ARCHITECTURE AND DIRECTIONAL ASSOCIATIONS BETWEEN ATTENTION-DEFICIT/HYPERACTIVITY AND BORDERLINE PERSONALITY DISORDERS: AN INTEGRATIVE GENOMIC ANALYSIS"

**Analysis of shared genetic architecture**

*Genome-wide genetic correlation and polygenic enrichment between phenotype pairs*

We applied linkage disequilibrium score regression (LDSC) to estimate the genome-wide genetic correlation between ADHD and BPD. This analysis can provide insight into the genetic relationship between these traits. To assess polygenic enrichment between phenotype pairs, we examined conditional quantile–quantile plots, which display the p-value distribution for one phenotype conditional on p-value thresholds for another. In a Q–Q plot, the y-axis represents the nominal p-value (p), whereas the x-axis represents the corresponding empirical cumulative distribution function value (q). Under the global null hypothesis, p-values are expected to follow a uniform distribution over the interval [0,1], producing a diagonal line when plotted against their empirical distribution. Leftward deviations of the observed distribution from this null expectation indicate increased tail probabilities of the test-statistic (z-score) distribution and, consequently, an excess of low p-values relative to chance expectations, referred to as enrichment. To emphasize differences in the distribution tails, Q–Q plots commonly display log10(p) against −log10(q). We generated conditional Q–Q plots showing the empirical quantiles of nominal −log10 p-values for SNP associations, both across all SNPs and within SNP subsets (strata) defined by the nominal p-values for their associations with the conditional phenotypes, and vice versa. Specifically, for a given phenotype, we estimated the empirical cumulative distribution of nominal p-values among all SNPs and among SNPs meeting the specified significance thresholds for the conditional phenotypes (−log10(p) ≥ 1, −log10(p) ≥ 2,

-log10(p) ≥ 3, corresponding to p < 0.1, p < 0.01, and p < 0.001, respectively). Nominal p-values, expressed as −log10(p), were plotted on the y-axis, whereas empirical quantiles, expressed as −log10(q), where q = 1 − cdf(p), were plotted on the x-axis. To limit potential inflation, the major histocompatibility complex (MHC) region was excluded was excluded.

*Identification of shared loci*

Shared loci were identified using conjFDR analysis. This approach consists of two conditional FDR (condFDR) analyses: in the first, associations with a secondary phenotype are used to re-rank test statistics for the primary phenotype; in the second, the phenotype roles are reversed. FDR may be understood as the probability that a SNP is null, conditional on its p-value being equal to or smaller than the observed value. The conditional FDR (condFDR) extends the conventional FDR framework by incorporating GWAS summary-statistic information from a second phenotype to adjust the significance assigned to a SNP. Specifically, condFDR represents the probability that a SNP is null for the primary phenotype, given that the p-values observed for both the primary and secondary phenotypes are at least as small as those observed. condFDR estimates are calculated for each nominal SNP p-value in the primary phenotype by first deriving stratified empirical cumulative distribution functions (cdfs) of nominal p-values (Karadag et al., 2023). These strata are defined according to the relative enrichment of SNP associations at progressively stronger nominal SNP associations with the secondary phenotype.

The standard FDR approach is based on a mixture model in which GWAS test statistics arise from both null and non-null effects, with true associations expected, on average, to yield more extreme test statistics than false associations. Consequently, ranking SNPs by standard FDR produces the same ordering as ranking them by p-value. However, when the primary and secondary phenotypes share genetic associations, condFDR can reorder SNPs, producing a ranking that differs from one based solely on p-values. Conjunctional FDR (conjFDR) refers to the posterior probability that a SNP is null for one phenotype, the other phenotype, or both, conditional on its association p-values for both phenotypes being equal to or smaller than the observed values (Karadag et al., 2023). A conservative conjFDR estimate is obtained by calculating condFDR in both trait directions and selecting the higher condFDR value for each SNP. Variants lacking summary statistics in either discovery dataset could not be evaluated for overlapping associations using conjFDR analysis. The higher of the two condFDR values is assigned as the conjFDR value. To mitigate the effects of linkage disequilibrium while retaining genome-wide information, the condFDR/conjFDR procedure incorporates repeated random LD pruning. For the primary analyses, 500 random pruning iterations were conducted for ADHD-BPD pair. To ensure reproducibility, the MATLAB random number generator was initialized with a fixed seed of 123 before batch processing.According to previous reports (Smeland et al., 2020), SNPs with conjFDR<0.05 were classified as shared.

To evaluate the directional consistency of the loci identified through conjFDR analysis, we performed sign-concordance analyses (Karadag et al., 2023), using independent or overlap-reduced validation datasets. For this purpose, we used summary statistics from FinnGen Release 13 (R13), specifically F5_ADHD and F5_EMOPER, corresponding to attention-deficit/hyperactivity disorder (ADHD) and emotionally unstable personality disorder, respectively.

Inspection of the cohort composition of the BPD discovery GWAS by Streit et al. revealed sample overlap due to the inclusion of FinnGen Release 8 (R8) F5_EMOPER data. To address this overlap, and given the lack of alternative sufficiently independent BPD GWAS datasets, we used MetaSubtract to remove the FinnGen R8 F5_EMOPER contribution from the Streit et al. meta-analysis, generating an R8-depleted BPD GWAS for directional validation. The reconstructed dataset underwent comprehensive quality control across 5,075,674 variants, including verification of allelic harmonization, variant uniqueness, completeness and validity of summary statistics, and positivity of residual inverse-variance weights. Internal statistical consistency was assessed by recalculating P-values from effect estimates and standard errors. In addition, the MetaSubtract output was independently reproduced using inverse-variance subtraction with genomic-control adjustments, demonstrating near-perfect agreement in effect estimates and standard errors (median absolute differences of 6.08 × 10⁻⁶ and 1.96 × 10⁻⁶, respectively). Detailed quality-control results are provided in Supplementary Table 4.

Sign concordance was assessed separately for each phenotype by comparing: (i) ADHD effect estimates from the Demontis et al. discovery GWAS with those from FinnGen R13 F5_ADHD; and (ii) BPD effect estimates from the FinnGen R8-depleted Streit et al. GWAS with those from FinnGen R13 F5_EMOPER. After harmonization of effect alleles, we calculated the proportion of lead variants showing concordant effect directions according to the signs of their beta coefficients. Under the null hypothesis of no directional consistency, the expected concordance proportion was 0.5. Statistical significance was assessed using two-sided exact binomial tests. Loci were considered directionally concordant when concordant effect directions were observed in both phenotype-specific comparisons. Variants that were unavailable, could not be reliably harmonized, or showed discordant effects in either comparison were not considered jointly replicated.

*Gene Mapping and Enrichment Analyses*

To elucidate the biological mechanisms contributing to the shared genetic architecture of attention-deficit/hyperactivity disorder and borderline personality disorder, loci identified by conjunctional false discovery rate (conjFDR) analysis were functionally characterized and mapped to putative genes using Functional Mapping and Annotation of Genome-Wide Association Studies. These genes were then examined through functional enrichment analyses of Gene Ontology terms and Kyoto Encyclopedia of Genes and Genomes pathways, as well as analyses of tissue- and cell-type-specific expression.

**Mendelian randomization**

In all MR statistics, genetic instruments were selected from the corresponding GWAS based on genome-wide significance (p < 5 x 10⁻⁸). Variants were clumped using the European subset of the 1000 Genomes Project Phase 3 as the linkage disequilibrium (LD) reference, with an r² < 0.001 threshold applied within a 10-Mb window. Only variants demonstrating adequate strength (F > 10) were retained. The primary MR analysis used inverse-variance weighted random-effects models (IVW). To complement this approach, we used the weighted-median estimator, which can provide consistent estimates if up to approximately 50% of the total instrument weight comes from invalid instruments, subject to its underlying assumptions (Burgess et al., 2023). Additional methods such as MR-Egger, MR pleiotropy residual sum and outlier (MR-PRESSO; Verbanck et al.,2018) and radial-MR, served as supplementary techniques for estimating causal effects. In essence, MR-Egger identifies small study bias in meta-analysis by performing a weighted linear regression of outcome coefficients against exposure coefficients, providing a valid and consistent causal effect estimate even when all genetic variants are invalid IVs. Nonetheless, this method is significantly less efficient than IVW and median-based approaches and may lack precision if all genetic variants exert a similar influence on exposure (Bowden et al., 2016). The weighted median technique aggregates data from multiple genetic variants into a single causal estimate, permitting up to 50% of the information to originate from invalid IVs (Bowden et al., 2016). In addition, between-instrument heterogeneity was assessed using Cochran’s Q statistic and I² confidence intervals (Burgess et al., 2023). We also performed leave-one-out analyses (LOO), sequentially removing each genetic instrument and repeating the IVW to determine whether individual variants disproportionately influenced the association. To assess whether the genetic instruments supported the proposed direction of effect, we performed Steiger directionality testing across multiple plausible prevalence scenarios for ADHD and BPD, informed by previously reported prevalence estimates. This test evaluates whether the instruments explain a greater proportion of phenotypic variance in the designated exposure than in the outcome (Burgess et al., 2023). Finally, to assess the potential influence of sample overlap and weak-instrument bias on the bidirectional MR estimates, we additionally applied MRlap (Mounier & Kutalik, 2023). This method uses genome-wide summary statistics together with LDSC estimates to quantify and correct biases arising from sample overlap, winner’s curse, and weak instruments. LDSC was used to estimate SNP-based heritability and cross-trait intercepts, with the latter providing information on potential correlation between GWAS estimation errors, including that attributable to sample overlap. MRlap was performed separately for the ADHD-to-BPD and BPD-to-ADHD analyses, and the bias-corrected MR estimates were compared with the corresponding conventional IVW estimates to assess the magnitude and direction of potential bias. MRlap derived signed Z-statistics from association coefficients and standard errors and performed cross-trait LD score regression through GenomicSEM, using precomputed European 1000 Genomes LD scores, originally distributed by the Alkes Group, and obtained from Zenodo (Pain, 2023; doi:10.5281/zenodo.8182036). The same LD scores were used as regression predictors and weights.. Instruments were selected independently in each direction using the native MRlap procedure, with an exposure-association threshold of p < 5 × 10⁻⁸, distance-based pruning at 500 kb, and the default reverse-association filter (MR_reverse = 0.001). This yielded 26 instruments for ADHD → BPD (Supplementary Table 9) and five for BPD → ADHD (supplementary Table 10). LDSC-derived exposure heritability and the cross-trait intercept informed the correction, together with estimated genetic-architecture parameters and the package’s fixed marker-count parameter (M = 1,150,000). Observed and corrected MR effects were expressed in standardized units on the observed scale. The package’s adaptive simulation procedure estimated the standard error of the corrected effect and its covariance with the observed estimate, using 6,000 simulations for ADHD → BPD and 5,000 for BPD → ADHD in the execution. Differences between observed and bias-corrected estimates were assessed using a two-sided Z-test that accounted for their covariance, estimated through the package’s adaptive simulation procedure. A random seed of 20261105 was set separately for each direction, and input snapshots, selected instruments, package versions, reference-file checksums, and munging and LDSC logs were retained. For case-control GWAS, interpretation additionally relies on the MRlap assumption that sample overlap is independent of case-control status.

To assess whether the genetic instruments supported the proposed direction of effect, we performed Steiger directionality testing, which evaluates whether they explained more phenotypic variance in the exposure than in the outcome (Burgess et al., 2023). Because both traits were binary, SNP–phenotype correlations were estimated on the liability scale using effect estimates, allele frequencies, numbers of cases and controls, and assumed population prevalences. Primary prevalence values of 5% for ADHD and 1.5% for BPD were used. To assess the sensitivity of reverse-direction results to these assumptions, we repeated the Steiger analyses across combinations of ADHD prevalences ranging from 3% to 10% and BPD prevalences ranging from 0.5% to 3%.

Additionally, to assess the extent to which the bidirectional MR associations were dependent on genomic regions showing strong evidence of shared genetic architecture, MR analyses were repeated after excluding instruments located within strongly colocalized loci (see below). IVW and weighted-median estimates were recalculated after exclusion, and changes in effect magnitude and heterogeneity were compared with the primary analyses. This analysis was intended to determine whether the observed MR associations were entirely attributable to the principal shared loci or remained detectable after their removal.

**Causal Analysis Using Summary Effect estimates**

Causal Analysis Using Summary Effect estimates (CAUSE; Morrison et al., 2020). Unlike conventional MR sensitivity methods, CAUSE uses genome-wide summary statistics and explicitly models correlated pleiotropy by comparing a sharing model, in which a proportion of variants may affect both traits through a shared factor, with a causal model that additionally allows for a causal effect of the exposure on the outcome. After harmonization of the ADHD and BPD GWAS datasets, 4,729,831 variants were available for analysis. Variants associated with the exposure at p < 1 × 10⁻³ were LD-pruned at r² < 0.1 using the CAUSE LD reference panel. This resulted in 2,135 variants for the ADHD-to-BPD analysis and 1,619 variants for the BPD-to-ADHD analysis. Nuisance parameters were estimated separately in each direction using a random sample of 1,000,000 genome-wide variants. Model fit was compared using the difference in expected log pointwise predictive density (ΔELPD), with evidence favoring the causal over the sharing model interpreted as indicating that correlated horizontal pleiotropy alone was insufficient to account for the observed association.

**Colocalization Analysis**

We also investigated whether the MR findings could partly reflect shared local genetic architecture. To do so, Bayesian colocalization analysis was performed using the coloc framework (Giambartolomei et al., 2014). Colocalization was conducted for all genetic instruments in both directions using genomic windows extending ±500 kb around each lead SNP (1 Mb in total). Effect estimates and their variances, allele frequencies, and sample-size information were obtained from the corresponding GWAS summary statistics. Alleles were harmonized between ADHD and BPD before analysis. The standard approximate Bayes factor approach (coloc.abf) was used to estimate posterior probabilities for five mutually exclusive hypotheses: no association with either phenotype (H0), association with ADHD only (H1), association with BPD only (H2), association with both phenotypes through distinct causal variants (H3), or association with both phenotypes through a shared causal variant (H4).

Posterior probability for H4 (PP.H4) was used to quantify evidence consistent with a shared local genetic signal, while high PP.H3 was interpreted as evidence favouring distinct association signals within the same genomic region. According to previous reports, loci with PP.H4 > 0.80 were considered to show strong evidence of colocalization for subsequent sensitivity analyses (Haglund et al., 2025). Loci showing strong evidence of colocalization in the approximate Bayes factor analysis were further evaluated using Sum of Single Effects (SuSiE; Wallace, 2021). Signed SNP-by-SNP LD matrices were calculated from the European Phase 3 1000 Genomes reference panel using the same set and ordering of variants included in each locus. GWAS effect estimates were aligned to the allele orientation used to calculate the LD matrix before fine-mapping. SuSiE was run separately for ADHD and BPD to identify credible sets representing statistically supported association signals within each region.

**Minimum Detectable Effect Analysis**

We calculated the minimum detectable effect corresponding to 80% power at a two-sided significance threshold of α = 0.05. Calculations were performed on the log-odds-ratio scale using harmonized SNP–exposure effect estimates and SNP–outcome standard errors. Under the inverse-variance weighted (IVW) fixed-effects model, the expected standard error of the causal estimate was calculated as

$$\boldsymbol{S}\boldsymbol{E}_{\boldsymbol{FE}}\boldsymbol{=}\left( \sum_{\boldsymbol{j=1}}^{\boldsymbol{k}} \frac{{\hat{\boldsymbol{\beta}}}_{\boldsymbol{Xj}}^{\boldsymbol{2}}}{\boldsymbol{SE}\left( {\hat{\boldsymbol{\beta}}}_{\boldsymbol{Yj}} \right)^{\boldsymbol{2}}} \right)^{\boldsymbol{-}\frac{\boldsymbol{1}}{\boldsymbol{2}}}$$

where β̂ₓⱼ denotes the association of SNP j with the exposure, SE(β̂ᵧⱼ) denotes the standard error of its association with the outcome, and k is the number of instruments. The minimum detectable absolute log odds ratio at 80% power was calculated as (z₁₋α/₂ + z₀.₈₀) × SE, where z denotes the corresponding standard normal quantile and α is the two-sided significance level. The corresponding detectable odds ratios were obtained as exp (±minimum detectable absolute log odds ratio).

To account for observed between-instrument heterogeneity, we additionally calculated conditional minimum detectable effects (MDEs) using multiplicative random-effects IVW standard errors:

$$\boldsymbol{S}\boldsymbol{E}_{\boldsymbol{MRE}}\boldsymbol{=S}\boldsymbol{E}_{\boldsymbol{FE}}\boldsymbol{\times}\sqrt{\max\left[ \boldsymbol{1,}\frac{\boldsymbol{Q}}{\boldsymbol{k-1}} \right]}$$

Here, Q denotes Cochran’s heterogeneity statistic, and k is the number of instruments. These conditional MDEs describe the detectable effect magnitude given the observed between-instrument dispersion and the resulting precision of each analysis. They were not interpreted as prospective power estimates or as corrections for horizontal pleiotropy.

The conditional minimum detectable absolute log odds ratio was subsequently calculated by replacing the fixed-effects standard error with the multiplicative random-effects standard error:

$$\left| \boldsymbol{\beta}_{\text{MDE,MRE}} \right|\boldsymbol{=}\left( \boldsymbol{z}_{\boldsymbol{1-}\boldsymbol{\alpha}\boldsymbol{/}\boldsymbol{2}}\boldsymbol{+}\boldsymbol{z}_{\boldsymbol{0.80}} \right)\boldsymbol{\times}\boldsymbol{S}\boldsymbol{E}_{\text{IVW,MRE}}$$

The corresponding detectable odds ratios were obtained by exponentiating the positive and negative thresholds. The procedure was applied separately to both directions of the bidirectional MR analysis and repeated after outlier exclusion using the instruments retained in each sensitivity analysis.

**Data availability**

The GWAS summary statistics for attention-deficit/hyperactivity disorder (ADHD) and borderline personality disorder (BPD) were obtained from publicly available datasets of Psychiatric Genomic consortium, associated with Demontis et al. (2023; https://doi.org/10.6084/m9.figshare.22564390) and Streit et al. (2026; https://doi.org/10.6084/m9.figshare.30777305). The FinnGen GWAS summary statistics used in this study are publicly available through the FinnGen data access portal (https://www.finngen.fi/en/access_results), including releases R13 and R8. Access to the summary statistics requires completion of the FinnGen online access form. No individual-level FinnGen participant data were accessed.

**Package software and statistical thresholds**

Statistical analyses were performed using R version 4.5.3, MATLAB R2026b (MathWorks, Natick, MA, USA), and established computational tools. Cross-trait enrichment and shared-locus discovery were conducted using the pleioFDR framework implemented in MATLAB, including conditional false discovery rate (condFDR) and conjunctional false discovery rate (conjFDR) analyses. SNP-based heritability and genome-wide genetic correlation were estimated using linkage disequilibrium score regression (LDSC), following the methods described by Bulik-Sullivan et al. (2015a, 2015b). Functional annotation, gene mapping, and gene-set enrichment analyses were performed using the Functional Mapping and Annotation (FUMA) platform.

Conventional bidirectional Mendelian randomization (MR) analyses were performed using MendelianRandomization (v0.10.0), TwoSampleMR (v0.7.0), and MRPRESSO (v1.0). Additional sensitivity analyses were conducted using RadialMR, MR-LAP to account for sample overlap and winner's curse, and CAUSE to distinguish causal effects from correlated horizontal pleiotropy. Colocalization analyses were performed using coloc (v5.2.3), and fine-mapping was conducted using susieR (v0.15.56). Data processing was performed using data.table (v1.18.4). Linkage disequilibrium-based clumping and LD calculations were performed using PLINK v2.0.0-b.1, with the 1000 Genomes Project Phase 3 European population as the LD reference panel.

Genome-wide significance was defined as p < 5 × 10⁻⁸ for genetic instrument selection. Statistical significance for conventional MR analyses was defined as a two-sided p < 0.05. The MR-PRESSO global test and Steiger directionality test were evaluated at p < 0.05. For colocalization analyses, posterior probabilities rather than p-value thresholds were used, with PP.H4 > 0.80 considered strong evidence of colocalization.

| **Cohort** | **Cases** | **Controls¹** | **Female cases** | **Female controls¹** | **Age group of cases** | **Design** | **Covariates** |
| --- | --- | --- | --- | --- | --- | --- | --- |
| iPSYCH1 | 17,019 | 21,880 | 28.2% | 49.5% | Children and adults | Case-control | PCs |
| iPSYCH2 | 8,876 | 15,268 | 35.5% | 49.8% | Children and adults | Case-control | PCs |
| deCODE | 8,281 | 137,993 | 47.0% | 54.6% | Children and adults | Case-control | SEX, Age, County |
| Barcelona, Spain | 572 | 425 | 30.2% | 23.5% | Adults | Case-control | PCs |
| Bergen, Norway | 295 | 202 | 53.6% | 60.9% | Adults | Case-control | PCs |
| Cardiff, UK | 721 | 5,081 | 12.9% | 49.4% | Children | Case-control | PCs |
| CHOP, USA | 262 | 262 | 24.4% | 24.4% | Children | Trios | None |
| Germany | 487 | 1,290 | 19.3% | 49.1% | Children | Case-control | PCs |
| IMAGE-I | 700 | 700 | 12.1% | 12.1% | Children | Trios | Site |
| IMAGE-II | 624 | 1,755 | 18.6% | 50.0% | Children | Case-control | PCs |
| PUWMa | 563 | 563 | 35.9% | 35.9% | Children | Trios | PCs |
| Toronto, Canada | 109 | 109 | 24.8% | 24.8% | Children | Trios | None |
| Yale-Penn | 182 | 1,315 | 30.2% | 42.2% | Adults | Case-control | PCs, alcohol dependence diagnosis |
| Total | 38,691 | 186,843 | — | — | Mixed | Mixed | Cohort-specific |

**SUPPLEMENTARY DATA**

**Supplementary Table 1.** Characteristics of cohorts contributing to the ADHD GWAS meta-analysis by Demontis et al. (2023).

| **Dataset** | **Cases** | **Controls** | **Female cases** | **Female controls** | **Age or birth year (SD), cases** | **Age or birth year (SD), controls** |
| --- | --- | --- | --- | --- | --- | --- |
| Germany | 993 | 1,539 | 91.5% | 56.2% | 30 (9) | 44 (13) |
| Central Europe | 1,285 | 1,332 | 85.0% | 60.3% | 31 (12) | 31 (14) |
| Spain 1 | 306 | 759 | 65.7% | 77.9% | 31 (7) | 28 (8) |
| Norway 1 | 57 | 700 | 89.5% | 44.7% | 27 (5) | 33 (9) |
| Norway 2 | 64 | 270 | 85.9% | 54.4% | 29 (7) | 36 (10 |
| HUNT | 295 | 42,535 | 78.0% | 47.5% | 1965 (12) | 1948 (17) |
| Estonian Biobank | 351 | 1,755 | 69.2% | 69.2% | 41 (11) | 41 (11) |
| UK Biobank | 182 | 1,820 | 70.3% | 70.8% | 50 (7) | 50 (7) |
| deCODE | 294 | 259,612 | 82.0% | 49.5% | 1972 (14) | 1960 (25) |
| MoBa | 386 | 7,730 | 85.5% | 85.5% | 45 (6) | 47 (5) |
| BioVU | 315 | 3,150 | 87.6% | 87.6% | 39 (15) | 40 (15) |
| CHB-DBDS | 2,379 | 334,766 | 81.7% | 52.6% | 1976 (15) | 1960 (20) |
| Mayo Clinic Biobank | 293 | 46,196 | 73.6% | 58.4% | 53 (16) | 66 (16) |
| GLAD | 1,327 | 6,703 | 88.5% | 52.5% | 34 (12) | 55 (14) |
| FinnGen* | 3,812 | 332,850 | 75.4% | 55.5% | 46 (14) | 59 (18) |

**Supplementary Table 2**. Characteristics of datasets contributing to the full BPD discovery GWAS meta-analysis by Streit et al (2026). All participants were of European ancestry. The first five datasets pooled individual-level data from 17 studies (2,705 cases; 4,600 controls; effective half N 3,175). The remaining ten datasets provided summary statistics (9,634 cases; 1,037,117 controls; effective half N 18,470).

* FinnGen Release 8 (R8) F5_EMOPER data

| **Endpoint** | **Official phenotype label** | **Cases** | **GWAS controls** | **GWAS total** | **Female cases** | **Male cases** | **Median age at first recorded event (years)** |
| --- | --- | --- | --- | --- | --- | --- | --- |
| F5_ADHD | Disturbance of activity and attention | 5,559 | 489,493 | 495,052 | 3,614 (65.0%) | 1,945 | 31.13 |
| F5_EMOPER | Emotionally unstable personality disorder | 5,676 | 485,845 | 491,521 | 4,387 (77.3%) | 1,289 | 30.11 |

**Supplementary Table 3**. Sample sizes and phenotype characteristics of the FinnGen R13 endpoints used for directional validation.

| QC domain | Quality-control metric | Observed result | Interpretation |
| --- | --- | --- | --- |
| Dataset integrity | Variants evaluated | 5,075,674 | Full harmonized variant set |
|  | Duplicate variant identifiers | 0 | No duplicated markers |
|  | Missing Beta, SE or p | 0 | Complete essential summary statistics |
| Allele harmonization | Discordant allele assignments | 0 | Allelic consistency verified |
| Statistical validity | Non-positive residual weights | 0 | All retained variants had positive residual inverse-variance weights |
|  | Invalid SE or P-values | 0 | No invalid statistical values detected |
| P-value consistency | Median absolute difference in −log10 P | 3.89 × 10⁻¹⁶ | Near-exact agreement |
|  | Maximum absolute difference in −log10 P | 7.28 × 10⁻¹⁴ | Negligible numerical discrepancy |
| Genomic control | FinnGen R8 λ used in reconstruction | 1.11 | Weight adjustment |
|  | Approximate reconstructed GWAS λ | 1.129 | SE inflation adjustment examined |
| Numerical reproducibility | Beta correlation | ≈ 1 | Near-perfect agreement; rounded output |
|  | Median absolute Beta difference | 6.08 × 10⁻⁶ | Very small discrepancy |
|  | Maximum absolute Beta difference | 2.16 × 10⁻⁴ | Small maximum discrepancy |
|  | SE correlation | ≈ 1 | Near-perfect agreement; rounded output |
|  | Median absolute SE difference | 1.96 × 10⁻⁶ | Very small discrepancy |
|  | Maximum absolute SE difference | 3.48 × 10⁻⁵ | Small maximum discrepancy |
|  | Median SE ratio, independent / MetaSubtract | 0.9999165 | Very close to unity |

**Supplementary table 4. Quality control of the FinnGen R8-depleted BPD GWAS summary statistics**. Direct comparison of the original Streit borderline personal disorder GWAS with the FinnGen R8-depleted dataset showed a strong correlation between effect estimates (r = 0.853) and standard errors (r = 0.989). Following subtraction, standard errors increased for all 5,075,674 variants, with a median SE ratio of 1.291, consistent with the expected reduction in statistical information. Allelic effect directions remained concordant for 82.97% of variants. Importantly, all 864,173 sign reversals occurred among variants with P ≥ 1 × 10⁻⁵ in the original GWAS, whereas no sign reversals were observed among the 1,311 variants meeting suggestive or genome-wide significance thresholds (P < 1 × 10⁻⁵). These findings support the statistical plausibility and internal consistency of the reconstructed summary statistics.

| **SNP** | **Chr** | **Position (hg19)** | **EA** | **OA** | **EAF** | **OR ADHD** | **β ADHD** | **SE** | **p** | **F** |
| --- | --- | --- | --- | --- | --- | --- | --- | --- | --- | --- |
| rs549845 | 1 | 44076469 | G | A | 0.33 | 1.08 | 0.08 | 0.01 | 9.03 × 10^−15^ | 59.69 |
| rs4916723 | 5 | 87854395 | A | C | 0.57 | 0.92 | -0.09 | 0.01 | 9.48 × 10^−15^ | 60.13 |
| rs2582895 | 11 | 28602173 | C | A | 0.62 | 1.08 | 0.07 | 0.01 | 4.09 × 10^−14^ | 57.03 |
| rs77960 | 5 | 103964585 | G | A | 0.68 | 0.93 | -0.07 | 0.01 | 2.46 × 10^−13^ | 53.59 |
| rs6082363 | 20 | 21250843 | T | C | 0.29 | 1.07 | 0.07 | 0.01 | 4.38 × 10^−12^ | 48.45 |
| rs9969232 | 7 | 114158954 | G | A | 0.38 | 0.93 | -0.07 | 0.01 | 9.98 × 10^−12^ | 46.65 |
| rs17576773 | 4 | 112217523 | C | T | 0.88 | 1.10 | 0.10 | 0.02 | 1.63 × 10^−10^ | 40.67 |
| rs114142727 | 3 | 87015142 | C | G | 0.99 | 1.28 | 0.25 | 0.04 | 5.13 × 10^−10^ | 38.67 |
| rs2886697 | 3 | 20724204 | G | A | 0.64 | 1.06 | 0.06 | 0.01 | 7.90 × 10^−10^ | 37.51 |
| rs76284431 | 14 | 98690923 | T | A | 0.84 | 0.92 | -0.08 | 0.01 | 1.19 × 10^−9^ | 37.08 |
| rs1162202 | 16 | 61966703 | C | T | 0.61 | 1.06 | 0.06 | 0.01 | 1.92 × 10^−9^ | 36.23 |
| rs704061 | 12 | 89771903 | T | C | 0.56 | 0.95 | -0.06 | 0.01 | 2.30 × 10^−9^ | 35.37 |
| rs17718444 | 3 | 71499401 | C | T | 0.67 | 1.06 | 0.06 | 0.01 | 2.87 × 10^−9^ | 35.30 |
| rs2025286 | 6 | 70858701 | A | C | 0.55 | 0.95 | -0.05 | 0.01 | 4.00 × 10^−9^ | 34.85 |
| rs1438898 | 2 | 145714354 | A | C | 0.77 | 1.06 | 0.06 | 0.01 | 4.88 × 10^−9^ | 33.92 |
| rs10875612 | 5 | 144474779 | C | T | 0.47 | 0.95 | -0.05 | 0.01 | 5.62 × 10^−9^ | 34.09 |
| rs9877066 | 3 | 43691501 | G | A | 0.95 | 0.89 | -0.12 | 0.02 | 6.60 × 10^−9^ | 33.63 |
| rs7844069 | 8 | 93277087 | T | G | 0.40 | 1.06 | 0.06 | 0.01 | 6.74 × 10^−9^ | 33.76 |
| rs4925811 | 8 | 145802447 | T | G | 0.53 | 0.94 | -0.06 | 0.01 | 8.30 × 10^−9^ | 32.98 |
| rs76857496 | 18 | 5871800 | C | A | 0.86 | 1.08 | 0.08 | 0.01 | 1.24 × 10^−8^ | 32.66 |
| rs7506904 | 18 | 50625779 | G | A | 0.37 | 0.95 | -0.06 | 0.01 | 1.24 × 10^−8^ | 32.54 |
| rs6537401 | 4 | 147099654 | G | A | 0.66 | 0.94 | -0.06 | 0.01 | 1.40 × 10^−8^ | 32.27 |
| rs11596214 | 10 | 106453832 | G | A | 0.57 | 1.05 | 0.05 | 0.01 | 3.17 × 10^−8^ | 30.89 |
| rs7613360 | 3 | 49916710 | C | T | 0.61 | 0.95 | -0.05 | 0.01 | 3.18 × 10^−8^ | 30.31 |
| rs73145587 | 7 | 67685754 | A | T | 0.90 | 1.11 | 0.10 | 0.02 | 3.67 × 10^−8^ | 30.31 |
| rs11255890 | 10 | 8784773 | C | A | 0.40 | 1.05 | 0.05 | 0.01 | 4.14 × 10^−8^ | 29.85 |

**Supplementary Table 5. Independent genome-wide significant genetic instruments for ADHD.** Independent variants were selected from the ADHD GWAS at genome-wide significance (p < 5 × 10−8) and LD-clumped using an r² threshold <0.001 within a 10-Mb window. OR and β are reported with respect to the effect allele (EA) in the ADHD GWAS. F statistics were calculated as (β/SE)². Abbreviations: ADHD, attention-deficit/hyperactivity disorder; EA, effect allele; OA, other allele; EAF, effect allele frequency; OR, odds ratio; SE, standard error; LD, linkage disequilibrium.

| **Exposure SNP** | **CHR** | **BP (hg19)** | **EA** | **OA** | **EAF ADHD** | **β ADHD** | **SE ADHD** | **p ADHD** | **F** | **β BPD** | **SE BPD** | **p BPD** |
| --- | --- | --- | --- | --- | --- | --- | --- | --- | --- | --- | --- | --- |
| rs549845 | 1 | 44076469 | G | A | 0.33 | 0.08 | 0.01 | 9.03x10^-15^ | 59.69 | 0.03 | 0.02 | 3.96x10^-02^ |
| rs2582895 | 11 | 28602173 | C | A | 0.62 | 0.07 | 0.01 | 4.09x10^-14^ | 57.03 | 0.06 | 0.01 | 3.98x10^-05^ |
| rs6082363 | 20 | 21250843 | T | C | 0.29 | 0.07 | 0.01 | 4.38x10^-12^ | 48.45 | 0.04 | 0.02 | 5.92x10^-03^ |
| rs9969232 | 7 | 114158954 | G | A | 0.38 | -0.07 | 0.01 | 9.98x10^-12^ | 46.65 | -0.09 | 0.02 | 1.99x10^-09^ |
| rs17576773 | 4 | 112217523 | C | T | 0.88 | 0.10 | 0.02 | 1.63x10^-10^ | 40.67 | 0.08 | 0.02 | 8.55x10^-04^ |
| rs114142727 | 3 | 87015142 | C | G | 0.99 | 0.25 | 0.04 | 5.13x10^-10^ | 38.67 | 0.13 | 0.06 | 2.29x10^-02^ |
| rs2886697 | 3 | 20724204 | G | A | 0.64 | 0.06 | 0.01 | 7.90x10^-10^ | 37.51 | -0.01 | 0.01 | 7.24x10^-01^ |
| rs76284431 | 14 | 98690923 | T | A | 0.84 | -0.08 | 0.01 | 1.19x10^-09^ | 37.08 | -0.02 | 0.02 | 3.78x10^-01^ |
| rs704061 | 12 | 89771903 | T | C | 0.56 | -0.06 | 0.01 | 2.30x10^-09^ | 35.37 | -0.04 | 0.01 | 1.07x10^-02^ |
| rs17718444 | 3 | 71499401 | C | T | 0.67 | 0.06 | 0.01 | 2.87x10^-09^ | 35.30 | 0.04 | 0.02 | 1.78x10^-02^ |
| rs2025286 | 6 | 70858701 | A | C | 0.55 | -0.05 | 0.01 | 4.00x10^-09^ | 34.85 | 0.00 | 0.01 | 8.69x10^-01^ |
| rs1438898 | 2 | 145714354 | A | C | 0.77 | 0.06 | 0.01 | 4.88x10^-09^ | 33.92 | 0.05 | 0.02 | 3.99x10^-03^ |
| rs10875612 | 5 | 144474779 | C | T | 0.47 | -0.05 | 0.01 | 5.62x10^-09^ | 34.09 | 0.01 | 0.01 | 5.29x10^-01^ |
| rs9877066 | 3 | 43691501 | G | A | 0.95 | -0.12 | 0.02 | 6.60x10^-09^ | 33.63 | 0.01 | 0.03 | 7.93x10^-01^ |
| rs7844069 | 8 | 93277087 | T | G | 0.40 | 0.06 | 0.01 | 6.74x10^-09^ | 33.76 | 0.02 | 0.01 | 2.49x10^-01^ |
| rs4925811 | 8 | 145802447 | T | G | 0.53 | -0.06 | 0.01 | 8.30x10^-09^ | 32.98 | -0.04 | 0.02 | 4.13x10^-03^ |
| rs76857496 | 18 | 5871800 | C | A | 0.86 | 0.08 | 0.01 | 1.24x10^-08^ | 32.66 | 0.00 | 0.02 | 9.69x10^-01^ |
| rs7506904 | 18 | 50625779 | G | A | 0.37 | -0.06 | 0.01 | 1.24x10^-08^ | 32.54 | -0.04 | 0.02 | 8.53x10^-03^ |
| rs6537401 | 4 | 147099654 | G | A | 0.66 | -0.06 | 0.01 | 1.40x10^-08^ | 32.27 | -0.01 | 0.02 | 4.31x10^-01^ |
| rs11596214 | 10 | 106453832 | G | A | 0.57 | 0.05 | 0.01 | 3.17x10^-08^ | 30.89 | 0.04 | 0.01 | 4.68x10^-03^ |
| rs7613360 | 3 | 49916710 | C | T | 0.61 | -0.05 | 0.01 | 3.18x10^-08^ | 30.31 | -0.04 | 0.02 | 1.28x10^-02^ |
| rs11255890 | 10 | 8784773 | C | A | 0.40 | 0.05 | 0.01 | 4.14x10^-08^ | 29.85 | 0.02 | 0.02 | 1.36x10^-01^ |

**Supplementary Table 6.** Genetic instruments included in the primary Mendelian randomization analysis of ADHD liability on BPD. The table reports the 22 genetic instruments included in the primary ADHD-to-BPD analysis after harmonization with the full BPD GWAS dataset. Effect estimates are expressed on the log-odds scale.

**Abbreviations:** ADHD, attention-deficit/hyperactivity disorder; BPD, borderline personality disorder; EAF, effect allele frequency; SE, standard error; LD, linkage disequilibrium. EA, effect allele; OA, other allele.

| **SNP** | **Chr** | **Position (hg19)** | **EA** | **OA** | **EAF** | **OR BPD** | **β BPD** | **SE** | **P** | **F** |
| --- | --- | --- | --- | --- | --- | --- | --- | --- | --- | --- |
| rs7304862 | 12 | 109,988,891 | G | T | 0.513 | 1.092 | 0.0879 | 0.0142 | 5.71 × 10⁻¹⁰ | 38.32 |
| rs4727799 | 7 | 114,110,568 | T | C | 0.670 | 1.098 | 0.0934 | 0.0151 | 6.92 × 10⁻¹⁰ | 38.26 |
| rs73581580 | 9 | 140,251,458 | G | A | 0.839 | 0.888 | −0.1187 | 0.0193 | 8.09 × 10⁻¹⁰ | 37.83 |
| rs2135029 | 5 | 155,856,538 | A | G | 0.599 | 0.922 | −0.0813 | 0.0143 | 1.41 × 10⁻⁸ | 32.32 |
| rs6549383 | 3 | 71,266,885 | T | C | 0.503 | 1.087 | 0.0832 | 0.0147 | 1.53 × 10⁻⁸ | 32.03 |
| rs9970854 | 1 | 191,125,935 | G | A | 0.597 | 0.922 | −0.0816 | 0.0145 | 1.99 × 10⁻⁸ | 31.67 |

**Supplementary Table 7. Independent genome-wide significant genetic instruments for borderline personality disorder.** Independent variants were selected from the BPD GWAS at genome-wide significance (P < 5 × 10⁻⁸) and LD-clumped using an r² threshold <0.001 within a 10-Mb window, using the 1000 Genomes Project Phase 3 European reference panel. OR and β are reported with respect to the BPD effect allele (EA). F statistics were calculated as (β/SE)². EA, effect allele; OA, other allele; EAF, effect allele frequency; OR, odds ratio; SE, standard error; BPD, borderline personality disorder; LD, linkage disequilibrium.

| **SNP** | **Chr** | **Position (hg19)** | **EA** | **OA** | **EAF BPD** | **β BPD** | **SE BPD** | **p BPD** | **EAF ADHD** | **β ADHD** | **SE ADHD** | **p ADHD** |
| --- | --- | --- | --- | --- | --- | --- | --- | --- | --- | --- | --- | --- |
| rs9970854 | 1 | 191125935 | G | A | 0.597 | −0.0816 | 0.0145 | 1.99×10⁻⁸ | 0.612 | −0.0254 | 0.0095 | 7.45×10⁻³ |
| rs6549383 | 3 | 71266885 | T | C | 0.503 | 0.0832 | 0.0147 | 1.53×10⁻⁸ | 0.516 | 0.0019 | 0.0097 | 8.48 ×10⁻^1^ |
| rs2135029 | 5 | 155856538 | A | G | 0.599 | −0.0813 | 0.0143 | 1.41×10⁻⁸ | 0.622 | −0.0210 | 0.0094 | 2.58×10⁻² |
| rs4727799 | 7 | 114110568 | T | C | 0.670 | 0.0934 | 0.0151 | 6.92×10⁻¹⁰ | 0.624 | 0.0641 | 0.0099 | 8.91×10⁻¹¹ |
| rs7304862 | 12 | 109988891 | G | T | 0.513 | 0.0879 | 0.0142 | 5.71×10⁻¹⁰ | 0.540 | 0.0164 | 0.0092 | 7.62×10⁻² |

**Supplementary Table 8. Genetic instruments included in the BPD-to-ADHD Mendelian randomization analysis.** Independent BPD-associated variants were selected at genome-wide significance (P < 5 × 10⁻⁸) and linkage disequilibrium (LD)-clumped at r² < 0.001 within a 10-Mb window using the 1000 Genomes Project Phase 3 European reference panel. Six independent BPD instruments were initially identified. One variant (rs73581580) was unavailable in the ADHD GWAS, and no proxy with r² ≥ 0.80 within ±1 Mb was available in the outcome dataset; therefore, five instruments were included in the BPD-to-ADHD analysis. ADHD effect estimates and effect allele frequencies were harmonized to the BPD effect allele shown in the table. F statistics refer to instrument strength for the BPD exposure and were calculated as (β/SE)². Abbreviations: ADHD, attention-deficit/hyperactivity disorder; BPD, borderline personality disorder; EA, effect allele; EAF, effect allele frequency; GWAS, genome-wide association study; LD, linkage disequilibrium; OA, other allele; SE, standard error.

| **MR-LAP SNP** | **Conventional MR SNP** | **Distance (kb)** | **LD r²** | **Classification** |
| --- | --- | --- | --- | --- |
| rs4131786 | rs1162202 | 186.145 | 0.3357810 | LD < 0.80 |
| rs6082363 | rs6082363 | 0.000 | 1.0000000 | Identical |
| rs3001723 | rs549845 | 38.784 | 0.9033560 | LD ≥ 0.80 |
| rs4925811 | rs4925811 | 0.000 | 1.0000000 | Identical |
| rs2886697 | rs2886697 | 0.000 | 1.0000000 | Identical |
| rs10795678 | rs11255890 | 28.856 | 0.9921030 | LD ≥ 0.80 |
| rs6537401 | rs6537401 | 0.000 | 1.0000000 | Identical |
| rs2582895 | rs2582895 | 0.000 | 1.0000000 | Identical |
| rs56331412 | rs17576773 | 1.350 | 1.0000000 | LD ≥ 0.80 |
| rs114142727 | rs114142727 | 0.000 | 1.0000000 | Identical |
| rs9877066 | rs9877066 | 0.000 | 1.0000000 | Identical |
| rs7613360 | rs7613360 | 0.000 | 1.0000000 | Identical |
| rs17718444 | rs17718444 | 0.000 | 1.0000000 | Identical |
| rs72854462 | rs1438898 | 5.785 | 1.0000000 | LD ≥ 0.80 |
| rs12819667 | rs704061 | 8.374 | 0.9962560 | LD ≥ 0.80 |
| rs11596214 | rs11596214 | 0.000 | 1.0000000 | Identical |
| rs2025286 | rs2025286 | 0.000 | 1.0000000 | Identical |
| rs30266 | rs77960 | 7.772 | 1.0000000 | LD ≥ 0.80 |
| rs76284431 | rs76284431 | 0.000 | 1.0000000 | Identical |
| rs2311059 | rs7613360 | 1967.362 | 0.0191205 | LD < 0.80 |
| rs10875612 | rs10875612 | 0.000 | 1.0000000 | Identical |
| rs7844069 | rs7844069 | 0.000 | 1.0000000 | Identical |
| rs76857496 | rs76857496 | 0.000 | 1.0000000 | Identical |
| rs12653396 | rs4916723 | 7.122 | 0.5536460 | LD < 0.80 |
| rs9969232 | rs9969232 | 0.000 | 1.0000000 | Identical |
| rs7506904 | rs7506904 | 0.000 | 1.0000000 | Identical |

**Supplementary Table 9. Instrument-level concordance: ADHD → BPD.** Individual SNP-level comparison of instruments selected by conventional LD-clumped MR and MR-LAP. Note. Pairwise LD was estimated using PLINK2 (--r2-unphased square) and the 1000 Genomes European reference panel, after matching genomic coordinates and alleles (GRCh37/hg19). Distance denotes absolute physical separation in kilobases. LD ≥ 0.80 denotes strong LD; identical SNPs have r² = 1.

| **MR-LAP SNP** | **Conventional MR SNP** | **Distance (kb)** | **LD r²** | **Classification** |
| --- | --- | --- | --- | --- |
| rs10850358 | rs7304862 | 2.159 | 1.000000 | LD ≥ 0.80 |
| rs6549383 | rs6549383 | 0.000 | 1.000000 | Identical |
| rs35624276 | rs4727799 | 70.465 | 0.546675 | LD < 0.80 |
| rs2135029 | rs2135029 | 0.000 | 1.000000 | Identical |
| rs9970854 | rs9970854 | 0.000 | 1.000000 | Identical |

**Supplementary Table 10. Instrument-level concordance: BPD → ADHD. Individual SNP-level comparison of instruments selected by conventional LD-clumped MR and MR-LAP.** Note. Pairwise LD was estimated using PLINK2 (--r2-unphased square) and the 1000 Genomes European reference panel, after matching genomic coordinates and alleles (GRCh37/hg19). Distance denotes absolute physical separation in kilobases. LD ≥ 0.80 denotes strong LD; identical SNPs have r² = 1.

| **BPD prevalence** | **ADHD prevalence** | **R² BPD** | **R² ADHD** | **Exposure /outcome R²** | **p-value** | **Consistent SNPs** |
| --- | --- | --- | --- | --- | --- | --- |
| 0.5% | 3% | 0.002131750 | 0.005594402 | 2.6243 | 1.49716 ×10^-4^ | 21/22 |
| 0.5% | 5% | 0.002131750 | 0.005593501 | 2.6239 | 1.50199×10^-4^ | 21/22 |
| 0.5% | 7% | 0.002131750 | 0.005592597 | 2.6235 | 1.50685×10^-4^ | 21/22 |
| 0.5% | 10% | 0.002131750 | 0.005591233 | 2.6228 | 1.51421×10^-4^ | 21/22 |
| 1.0% | 3% | 0.002131536 | 0.005594402 | 2.6246 | 1.49532×10^-4^ | 21/22 |
| 1.0% | 5% | 0.002131536 | 0.005593501 | 2.6242 | 1.50014×10^-4^ | 21/22 |
| 1.0% | 7% | 0.002131536 | 0.005592597 | 2.6237 | 1.50500×10^-4^ | 21/22 |
| 1.0% | 10% | 0.002131536 | 0.005591233 | 2.6231 | 1.51235×10^-4^ | 21/22 |
| 1.5% | 3% | 0.002131323 | 0.005594402 | 2.6249 | 1.49348×10^-4^ | 21/22 |
| 1.5% | 5% | 0.002131323 | 0.005593501 | 2.6244 | 1.49829×10^-4^ | 21/22 |
| 1.5% | 7% | 0.002131323 | 0.005592597 | 2.6240 | 1.50314×10^-4^ | 21/22 |
| 1.5% | 10% | 0.002131323 | 0.005591233 | 2.6234 | 1.51048×10^-4^ | 21/22 |
| 2.0% | 3% | 0.002131109 | 0.005594402 | 2.6251 | 1.49164×10^-4^ | 21/22 |
| 2.0% | 5% | 0.002131109 | 0.005593501 | 2.6247 | 1.49645×10^-4^ | 21/22 |
| 2.0% | 7% | 0.002131109 | 0.005592597 | 2.6243 | 1.50129×10^-4^ | 21/22 |
| 2.0% | 10% | 0.002131109 | 0.005591233 | 2.6236 | 1.50862×10^-4^ | 21/22 |
| 3.0% | 3% | 0.002130681 | 0.005594402 | 2.6256 | 1.48796×10^-4^ | 21/22 |
| 3.0% | 5% | 0.002130681 | 0.005593501 | 2.6252 | 1.49276×10^-4^ | 21/22 |
| 3.0% | 7% | 0.002130681 | 0.005592597 | 2.6248 | 1.49759×10^-4^ | 21/22 |
| 3.0% | 10% | 0.002130681 | 0.005591233 | 2.6242 | 1.50491×10^-4^ | 21/22 |

**Supplementary Table 11.** Steiger directionality analyses for ADHD → BPD. Steiger directionality was evaluated across 20 combinations of assumed population prevalences for BPD (0.5%–3.0%) and ADHD (3%–10%). Across all scenarios, the genetic instruments explained more variance in ADHD than in BPD (ADHD R² = 0.00559; BPD R² = 0.00213; exposure-to-outcome R² ratio = 2.62). Directionality was concordant with ADHD-to-BPD for 21 of 22 SNPs across all scenarios, supporting the robustness of the inferred orientation to plausible prevalence assumptions.

| **BPD prevalence** | **ADHD prevalence** | **R² BPD** | **R² ADHD** | **Exposure /outcome R²** | **p-value** | **Consistent SNPs** |
| --- | --- | --- | --- | --- | --- | --- |
| 0.5% | 3% | 0.002667231 | 0.000392227 | 6.8002 | 2.61679×10^-5^ | 5/5 |
| 0.5% | 5% | 0.002667231 | 0.000392144 | 6.8017 | 2.61359×10^-5^ | 5/5 |
| 0.5% | 7% | 0.002667231 | 0.000392061 | 6.8031 | 2.61039×10^-5^ | 5/5 |
| 0.5% | 10% | 0.002667231 | 0.000391936 | 6.8053 | 2.60557×10^-5^ | 5/5 |
| 1.0% | 3% | 0.002667209 | 0.000392227 | 6.8002 | 2.61711e-05 | 5/5 |
| 1.0% | 5% | 0.002667209 | 0.000392144 | 6.8016 | 2.61391×10^-5^ | 5/5 |
| 1.0% | 7% | 0.002667209 | 0.000392061 | 6.8030 | 2.61071×10^-5^ | 5/5 |
| 1.0% | 10% | 0.002667209 | 0.000391936 | 6.8052 | 2.60589×10^-5^ | 5/5 |
| 1.5% | 3% | 0.002667187 | 0.000392227 | 6.8001 | 2.61743×10^-5^ | 5/5 |
| 1.5% | 5% | 0.002667187 | 0.000392144 | 6.8015 | 2.61423×10^-5^ | 5/5 |
| 1.5% | 7% | 0.002667187 | 0.000392061 | 6.8030 | 2.61103×10^-5^ | 5/5 |
| 1.5% | 10% | 0.002667187 | 0.000391936 | 6.8052 | 2.60621×10^-5^ | 5/5 |
| 2.0% | 3% | 0.002667165 | 0.000392227 | 6.8001 | 2.61776×10^-5^ | 5/5 |
| 2.0% | 5% | 0.002667165 | 0.000392144 | 6.8015 | 2.61456×10^-5^ | 5/5 |
| 2.0% | 7% | 0.002667165 | 0.000392061 | 6.8029 | 2.61135×10^-5^ | 5/5 |
| 2.0% | 10% | 0.002667165 | 0.000391936 | 6.8051 | 2.60653×10^-5^ | 5/5 |
| 3.0% | 3% | 0.002667121 | 0.000392227 | 6.7999 | 2.61842×10^-5^ | 5/5 |
| 3.0% | 5% | 0.002667121 | 0.000392144 | 6.8014 | 2.61522×10^-5^ | 5/5 |
| 3.0% | 7% | 0.002667121 | 0.000392061 | 6.8028 | 2.61201×10^-5^ | 5/5 |
| 3.0% | 10% | 0.002667121 | 0.000391936 | 6.8050 | 2.60719×10^-5^ | 5/5 |

**Supplementary Table 12 Steiger directionality analyses for BPD → ADHD direction** Steiger filtering consistently supported the BPD-to-ADHD orientation across all 20 combinations of assumed BPD and ADHD prevalences. The five BPD instruments explained approximately 6.8-fold more variance in BPD than in ADHD with all instruments meeting the Steiger directionality criterion. These findings were robust to the prevalence assumptions considered.

| Index SNP | Chr | Region (Mb) | SNPs | PP.H0 | PP.H1 | PP.H2 | PP.H3 | PP.H4 |
| --- | --- | --- | --- | --- | --- | --- | --- | --- |
| rs9969232 | 7 | 113.659–114.659 | 1.110 | 0.0000 | 0.0000 | 0.0000 | 0.0098 | 0.9902 |
| rs17576773 | 4 | 111.718–112.718 | 1.653 | 0.0000 | 0.0047 | 0.0000 | 0.0235 | 0.9718 |
| rs2582895 | 11 | 28.102–29.102 | 1.621 | 0.0000 | 0.0120 | 0.0000 | 0.1130 | 0.8749 |
| rs11596214 | 10 | 105.954–106.954 | 2.065 | 0.0016 | 0.2351 | 0.0010 | 0.1468 | 0.6156 |
| rs7506904 | 18 | 50.126–51.126 | 2.751 | 0.0005 | 0.2846 | 0.0003 | 0.1996 | 0.5150 |
| rs4925811 | 8 | 145.302–146.302 | 572 | 0.0024 | 0.5394 | 0.0004 | 0.0833 | 0.3745 |
| rs1438898 | 2 | 145.214–146.214 | 1.465 | 0.0008 | 0.5453 | 0.0001 | 0.0819 | 0.3719 |
| rs7613360 | 3 | 49.417–50.417 | 1.002 | 0.0051 | 0.6109 | 0.0011 | 0.1285 | 0.2544 |
| rs114142727 | 3 | 86.515–87.515 | 2.253 | 0.0003 | 0.7872 | 0.0000 | 0.0299 | 0.1826 |
| rs6082363 | 20 | 20.751–21.751 | 1.425 | 0.0000 | 0.5585 | 0.0000 | 0.2944 | 0.1471 |
| rs704061 | 12 | 89.272–90.272 | 1.415 | 0.0006 | 0.8094 | 0.0000 | 0.0499 | 0.1401 |
| rs9877066 | 3 | 43.192–44.192 | 1.536 | 0.0003 | 0.5886 | 0.0002 | 0.3519 | 0.0589 |
| rs549845 | 1 | 43.576–44.576 | 1.705 | 0.0000 | 0.5882 | 0.0000 | 0.3805 | 0.0312 |
| rs7844069 | 8 | 92.777–93.777 | 1.418 | 0.0015 | 0.8404 | 0.0003 | 0.1411 | 0.0167 |
| rs11255890 | 10 | 8.285–9.285 | 2.737 | 0.0020 | 0.9269 | 0.0001 | 0.0556 | 0.0154 |
| rs76857496 | 18 | 5.372–6.372 | 1.911 | 0.0050 | 0.9260 | 0.0003 | 0.0592 | 0.0094 |
| rs6537401 | 4 | 146.600–147.600 | 1.753 | 0.0007 | 0.9398 | 0.0000 | 0.0502 | 0.0092 |
| rs10875612 | 5 | 143.975–144.975 | 1.993 | 0.0009 | 0.8798 | 0.0001 | 0.1117 | 0.0074 |
| rs2886697 | 3 | 20.224–21.224 | 2.846 | 0.0001 | 0.8891 | 0.0000 | 0.1035 | 0.0072 |
| rs2025286 | 6 | 70.359–71.359 | 2.151 | 0.0005 | 0.9255 | 0.0000 | 0.0672 | 0.0068 |
| rs76284431 | 14 | 98.191–99.191 | 3.166 | 0.0000 | 0.0141 | 0.0003 | 0.9795 | 0.0061 |
| rs17718444 | 3 | 70.999–71.999 | 1.726 | 0.0000 | 0.0154 | 0.0004 | 0.9817 | 0.0024 |

**Supplementary Table 13. Colocalization analysis of attention-deficit/hyperactivity disorder associated loci with borderline personality disorder.** Colocalization analysis was performed using the approximate Bayes factor. Genomic regions were defined as ±500 kb windows centered on the lead SNPs selected as genetic instruments for the Mendelian randomization analysis of ADHD on BPD. Prior probabilities were set to p1 = 1 × 10⁻⁴. p2 = 1 × 10⁻⁴. and p12 = 1 × 10⁻⁵.

| **Index SNP** | **Chr** | **Region (Mb)** | **SNPs** | **PP.H3 (ABF)** | **PP.H4 (ABF)** | **PP.H3 (SuSiE)** | **PP.H4 (SuSiE)** |
| --- | --- | --- | --- | --- | --- | --- | --- |
| rs9969232 | 7 | 113.659–114.659 | 1.110 | 0.0098 | 0.9902 | 0.0112 | 0.9887 |
| rs17576773 | 4 | 111.718–112.718 | 1.653 | 0.0235 | 0.9718 | 0.0248 | 0.9720 |
| rs2582895 | 11 | 28.102–29.102 | 1.621 | 0.1130 | 0.8749 | — | — |
| rs11596214 | 10 | 105.954–106.954 | 2.065 | 0.1468 | 0.6156 | — | — |
| rs7506904 | 18 | 50.126–51.126 | 2.751 | 0.1996 | 0.5150 | — | — |
| rs4925811 | 8 | 145.302–146.302 | 572 | 0.0833 | 0.3745 | — | — |
| rs1438898 | 2 | 145.214–146.214 | 1.465 | 0.0819 | 0.3719 | — | — |
| rs7613360 | 3 | 49.417–50.417 | 1.002 | 0.1285 | 0.2544 | — | — |
| rs114142727 | 3 | 86.515–87.515 | 2.253 | 0.0299 | 0.1826 | — | — |
| rs6082363 | 20 | 20.751–21.751 | 1.425 | 0.2944 | 0.1471 | — | — |
| rs704061 | 12 | 89.272–90.272 | 1.415 | 0.0499 | 0.1401 | — | — |
| rs9877066 | 3 | 43.192–44.192 | 1.536 | 0.3519 | 0.0589 | — | — |
| rs549845 | 1 | 43.576–44.576 | 1.705 | 0.3805 | 0.0312 | — | — |
| rs7844069 | 8 | 92.777–93.777 | 1.418 | 0.1411 | 0.0167 | — | — |
| rs11255890 | 10 | 8.285–9.285 | 2.737 | 0.0556 | 0.0154 | — | — |
| rs76857496 | 18 | 5.372–6.372 | 1.911 | 0.0592 | 0.0094 | — | — |
| rs6537401 | 4 | 146.600–147.600 | 1.753 | 0.0502 | 0.0092 | — | — |
| rs10875612 | 5 | 143.975–144.975 | 1.993 | 0.1117 | 0.0074 | — | — |
| rs2886697 | 3 | 20.224–21.224 | 2.846 | 0.1035 | 0.0072 | — | — |
| rs2025286 | 6 | 70.359–71.359 | 2.151 | 0.0672 | 0.0068 | — | — |
| rs76284431 | 14 | 98.191–99.191 | 3.166 | 0.9795 | 0.0061 | 0.9761 | 0.0009 |
| rs17718444 | 3 | 70.999–71.999 | 1.726 | 0.9817 | 0.0024 | 0.9893 | 0.0007 |

**Supplementary Table 14. Comparison of coloc-ABF and coloc-SuSiE results for** **attention-deficit/hyperactivity disorder-associated loci and borderline personality disorder.** Coloc-ABF was applied to ±500 kb windows centered on 22 ADHD index SNPs (GRCh37/hg19). SuSiE-based colocalization posterior probabilities are shown only for the four loci with an evaluable signal pair; em dashes denote no evaluable coloc-SuSiE result. not evidence against colocalization. H3 indicates distinct causal variants and H4 indicates a shared causal variant. Prior probabilities were p1 = 1 × 10⁻⁴. p2 = 1 × 10⁻⁴. and p12 = 1 × 10⁻⁵.

| **Index SNP** | **Chr** | **Region (Mb)** | **SNPs** | **PP.H0** | **PP.H1** | **PP.H2** | **PP.H3** | **PP.H4** |
| --- | --- | --- | --- | --- | --- | --- | --- | --- |
| rs9970854 | 1 | 190.626–191.626 | 2.093 | 0.0006 | 0.0001 | 0.8265 | 0.1045 | 0.0684 |
| rs6549383 | 3 | 70.767–71.767 | 1.449 | 0.0000 | 0.0154 | 0.0004 | 0.9817 | 0.0024 |
| rs2135029 | 5 | 155.357–156.357 | 2.008 | 0.0019 | 0.0001 | 0.9148 | 0.0464 | 0.0367 |
| rs4727799 | 7 | 113.611–114.611 | 1.114 | 0.0000 | 0.0000 | 0.0000 | 0.0098 | 0.9902 |
| rs7304862 | 12 | 109.489–110.489 | 2.193 | 0.0000 | 0.0000 | 0.9331 | 0.0463 | 0.0206 |

**Supplementary Table 15. Colocalization analysis of borderline personality disorders-associated loci with attention-deficit/hyperactivity disorder** Colocalization analysis was performed using the approximate Bayes factor method. Genomic regions were defined as ±500 kb windows centered on the lead SNPs selected as genetic instruments for the Mendelian randomization analysis of BPD on ADHD. Prior probabilities were set to p1 = 1 × 10⁻⁴. p2 = 1 × 10⁻⁴. and p12 = 1 × 10⁻⁵.

| **Index SNP** | **Chr** | **Region (Mb)** | **SNPs** | **PP.H3 (ABF)** | **PP.H4 (ABF)** | **PP.H3 (SuSiE)** | **PP.H4 (SuSiE)** |
| --- | --- | --- | --- | --- | --- | --- | --- |
| rs4727799 | 7 | 113.611–114.611 | 1.114 | 0.0098 | 0.9902 | 0.0148 | 0.9851 |
| rs6549383 | 3 | 70.767–71.767 | 1.449 | 0.9817 | 0.0024 | 0.9886 | 0.0006 |
| rs9970854 | 1 | 190.626–191.626 | 2.093 | 0.1045 | 0.0684 | — | — |
| rs2135029 | 5 | 155.357–156.357 | 2.008 | 0.0464 | 0.0367 | — | — |
| rs7304862 | 12 | 109.489–110.489 | 2.193 | 0.0463 | 0.0206 | — | — |

**Supplementary Table 16. Comparison of coloc-ABF and coloc-SuSiE results for borderline personality disorder-associated loci and attention-deficit/hyperactivity disorder.** Coloc-ABF was applied to ±500 kb windows centered on five BPD index SNPs (GRCh37/hg19). SuSiE-based colocalization posterior probabilities are shown only for the two loci with an evaluable signal pair; em dashes denote no evaluable coloc-SuSiE result and are not evidence against colocalization. H3 indicates distinct causal variants, whereas H4 indicates a shared causal variant. Prior probabilities were to p1 = 1 × 10⁻⁴. p2 = 1 × 10⁻⁴. and p12 = 1 × 10⁻⁵.

| **Analysis** | **SNPs** | **β** | **SE** | **p-value** |
| --- | --- | --- | --- | --- |
| IVW, multiplicative random effects | 20 | 0.417222 | 0.074007 | 1.72 × 10⁻⁸ |
| Weighted median | 20 | 0.420407 | 0.088440 | 2.00 × 10⁻⁶ |
| MR-Egger | 20 | 0.335487 | 0.300503 | 2.79 × 10⁻^1^ |
| MR-PRESSO, uncorrected | 20 | 0.417222 | 0.074007 | 1.95 × 10⁻⁵ |
| Radial IVW, modified second-order | 20 | 0.417222 | 0.074007 | 1.74 × 10⁻⁸ |
| Radial IVW, iterative | 20 | 0.417222 | 0.074007 | 1.74 × 10⁻⁸ |
| Radial IVW, exact fixed effects | 20 | 0.4354 | 0.0581 | 6.42 × 10⁻¹⁴ |
| Radial IVW, exact random effects | 20 | 0.4282 | 0.0734 | 1.28 × 10⁻⁵ |
| Radial IVW, modified second-order | 18 | 0.411364 | 0.071144 | 7.38 × 10⁻⁹ |
| Radial IVW, iterative | 18 | 0.411364 | 0.071144 | 7.38 × 10⁻⁹ |
| Radial IVW, exact fixed effects | 18 | 0.43 | 0.06 | 5.70 × 10⁻¹² |
| Radial IVW, exact random effects | 18 | 0.42 | 0.07 | 7.52 × 10⁻⁶ |

**Supplementary Table 17. Sensitivity analyses of the association between genetic liability to attention-deficit/hyperactivity disorder and borderline personality disorder after exclusion of lead variants from colocalized loci.** Mendelian randomization analyses estimating the association between genetic liability to attention-deficit/hyperactivity disorder (ADHD) and liability to borderline personality disorder (BPD) following exclusion of lead single-nucleotide polymorphisms (SNPs) at loci showing evidence of colocalization between the two traits. The primary colocalization-informed sensitivity analysis excluded rs9969232 and rs17576773, leaving 20 independent genetic instruments. Exploratory radial Mendelian randomization analyses additionally excluded rs2582895 and rs10875612, leaving 18 instruments. Effect estimates (β) represent the estimated change in BPD liability per unit increase in genetically predicted ADHD liability.

**Abbreviations:** β, Mendelian randomization effect estimate; IVW, inverse-variance weighted; MR-Egger, Mendelian randomization Egger regression; MR-PRESSO, Mendelian Randomization Pleiotropy RESidual Sum and Outlier; SE, standard error; SNP, single-nucleotide polymorphism.

| **Diagnostic** | **Result** | **p-value** |
| --- | --- | --- |
| IVW heterogeneity (20 SNPs) | Q = 33.4257; df = 19; I² = 43.2% | 0.0215 |
| MR-Egger heterogeneity (20 SNPs) | Q = 33.3; df = 18 | 0.0153 |
| MR-Egger intercept | 0.005437 (SE = 0.019342) | 0.782 |
| MR-PRESSO global test | RSSobs = 37.06; no MR-PRESSO outliers | 0.0398 |
| Radial heterogeneity (20 SNPs) | Q = 31.09; df = 19; I² = 38.9% | 0.0393 |
| Radial outlier rs2582895 | Individual Q = 3.9 | 0.048 |
| Radial outlier rs10875612 | Individual Q = 4.6 | 0.032 |
| Radial heterogeneity (18 SNPs) | Q = 22.5909; df = 17; I² = 24.7%; no further nominal outliers | 0.163 |

**Supplementary Table 18. Heterogeneity and horizontal pleiotropy diagnostics for Mendelian randomization analysis of the association between genetic liability to attention-deficit/hyperactivity disorder and borderline personality disorder, after exclusion of lead variants from colocalized loci**. Moderate between-instrument heterogeneity was observed in the 20-SNP analysis. The MR-Egger intercept provided no evidence of directional horizontal pleiotropy. Although the MR-PRESSO global test indicated overall heterogeneity, no outlier variants were identified by MR-PRESSO. Radial Mendelian randomization identified rs2582895 and rs10875612 as nominal outliers; after their exclusion, heterogeneity was attenuated and no additional nominal radial outliers were detected.

**Abbreviations:** ADHD, attention-deficit/hyperactivity disorder; BPD, borderline personality disorder; df, degrees of freedom; I², proportion of variation in instrument-specific estimates attributable to heterogeneity; IVW, inverse-variance weighted; MR, Mendelian randomization; MR-Egger, Mendelian randomization Egger regression; MR-PRESSO, Mendelian Randomization Pleiotropy RESidual Sum and Outlier; Q, Cochran’s heterogeneity statistic; RSSobs, observed residual sum of squares; SE, standard error; SNP, single-nucleotide polymorphism.

| **Analysis** | **SNPs** | **β** | **SE** | **P-value** |
| --- | --- | --- | --- | --- |
| IVW, multiplicative random effects | 4 | 0.194613 | 0.060872 | 0.00139 |
| IVW, fixed effects | 4 | 0.194613 | 0.056484 | 0.000570 |
| Weighted median | 4 | 0.211356 | 0.072206 | 0.00342 |
| MR-Egger | 4 | −0.794440 | 2.209676 | 0.754 |
| Radial IVW, modified second-order | 4 | 0.194579 | 0.060900 | 0.0495 |
| Radial IVW, iterative | 4 | 0.1946 | 0.0609 | 0.0495 |
| Radial IVW, exact fixed effects | 4 | 0.199273 | 0.059043 | 0.0432 |
| Radial IVW, exact random effects | 4 | 0.198948 | 0.053028 | 0.0331 |

**Supplementary Table 19. Sensitivity analyses of the association between genetic liability to borderline personality disorder and attention-deficit/hyperactivity disorder after exclusion of lead variant** **rs4727799 from co-localized locus.** Across the four SNP instruments, inverse-variance weighted (IVW) analyses indicated a positive association (multiplicative random-effects β = 0.195, SE = 0.061, P = 0.00139), consistent with the weighted-median estimate (β = 0.211, SE = 0.072, P = 0.00342). Radial IVW estimates were similarly positive and nominally significant across model specifications. In contrast, the MR-Egger estimate was imprecise and non-significant (β = −0.794, SE = 2.210, P = 0.754), likely reflecting the limited number of instruments. For radial IVW analyses, p-values were calculated using t-distribution-based inference with residual degrees of freedom, accounting for the limited number of genetic instruments.

| **Diagnostic** | **Result** | **P-value** |
| --- | --- | --- |
| IVW heterogeneity | Q = 3.484309; df = 3; I² = 13.9% | 0.323 |
| MR-Egger heterogeneity | Q = 3.167; df = 2 | 0.205 |
| MR-Egger intercept | 0.082734 (SE = 0.184744) | 0.698 |
| MR-PRESSO global test | RSSobs = 5.959; no outliers | 0.4291 |
| Radial IVW heterogeneity | Q = 3.204; df = 3; I² = 6.4% | 0.361 |
| Radial outliers | None detected | — |

**Supplementary Table 20. Heterogeneity and horizontal pleiotropy diagnostics for Mendelian randomization (MR) analysis of the association between genetic liability to borderline personality disorder and attention-deficit/hyperactivity disorder after exclusion of lead variant** **of lead variant** **rs4727799 from co-localized locus.** . No between-instrument heterogeneity was observed in the 4-SNP analysis. The MR-Egger intercept provided no evidence of directional horizontal pleiotropy. No outlier variants were identified by MR-PRESSO and Radial MR. However, due the limited number of genetic instruments the data may be interpreted with caution

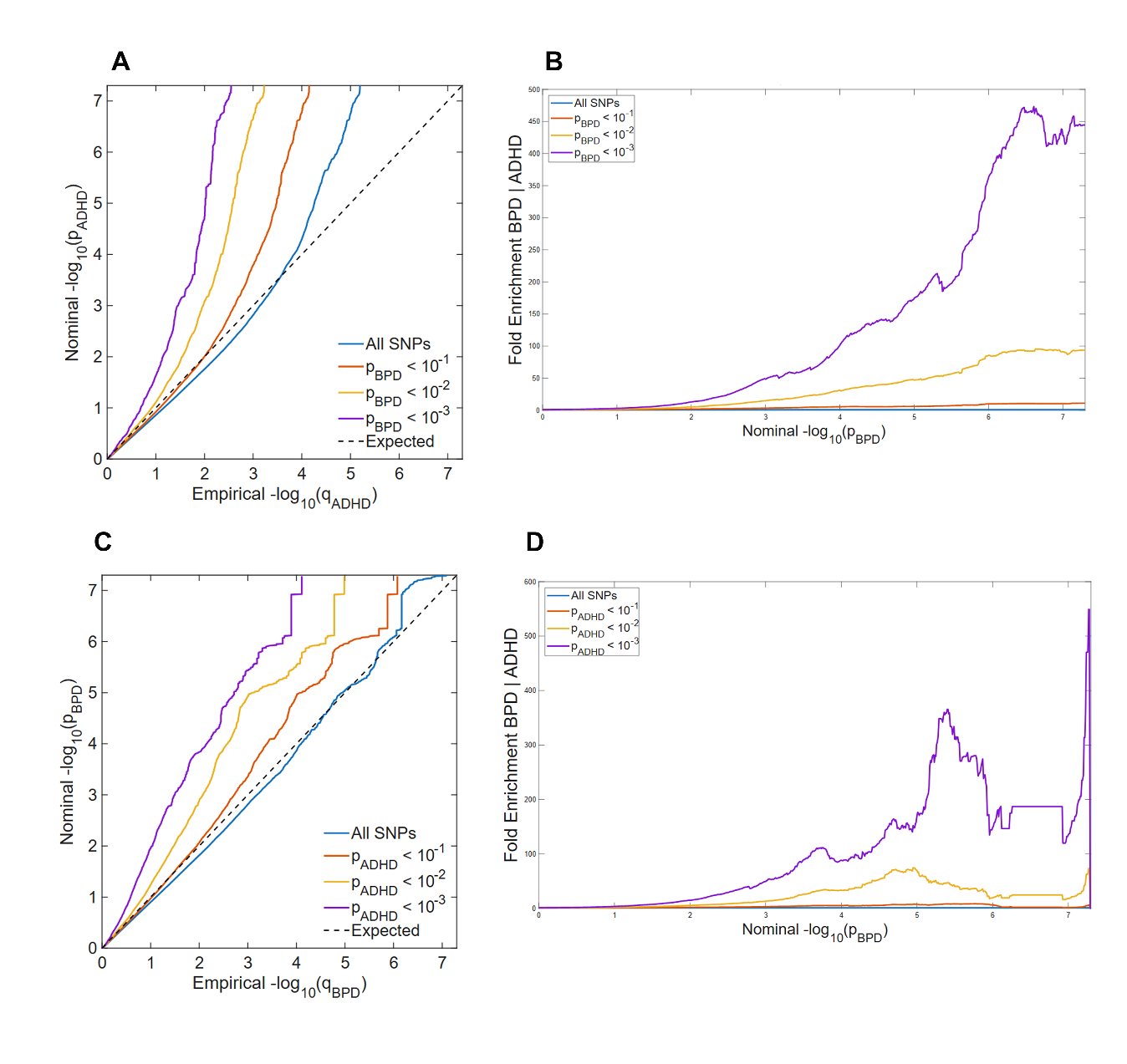

**Supplementary Figure 1. Cross-trait enrichment between attention-deficit/hyperactivity disorder (ADHD) and borderline personality disorder (BPD).** Conditional quantile–quantile (Q–Q) plots (A. C) and corresponding fold-enrichment plots (B. D) illustrate the enrichment of genetic associations with ADHD conditional on BPD (panels A and B) and with BPD conditional on ADHD (panels C and D). SNPs were stratified according to progressively stringent association thresholds with the conditioning phenotype (P < 10⁻¹. 10⁻². and 10⁻³). In the conditional Q–Q plots. increasing leftward deviations from the null expectation indicate progressively stronger cross-trait enrichment. Fold-enrichment plots show the corresponding increase in the proportion of SNPs exceeding nominal significance thresholds relative to the unconditioned distribution. Both directions demonstrate cross-trait enrichment. with a particularly pronounced enrichment of ADHD-associated variants conditional on BPD.

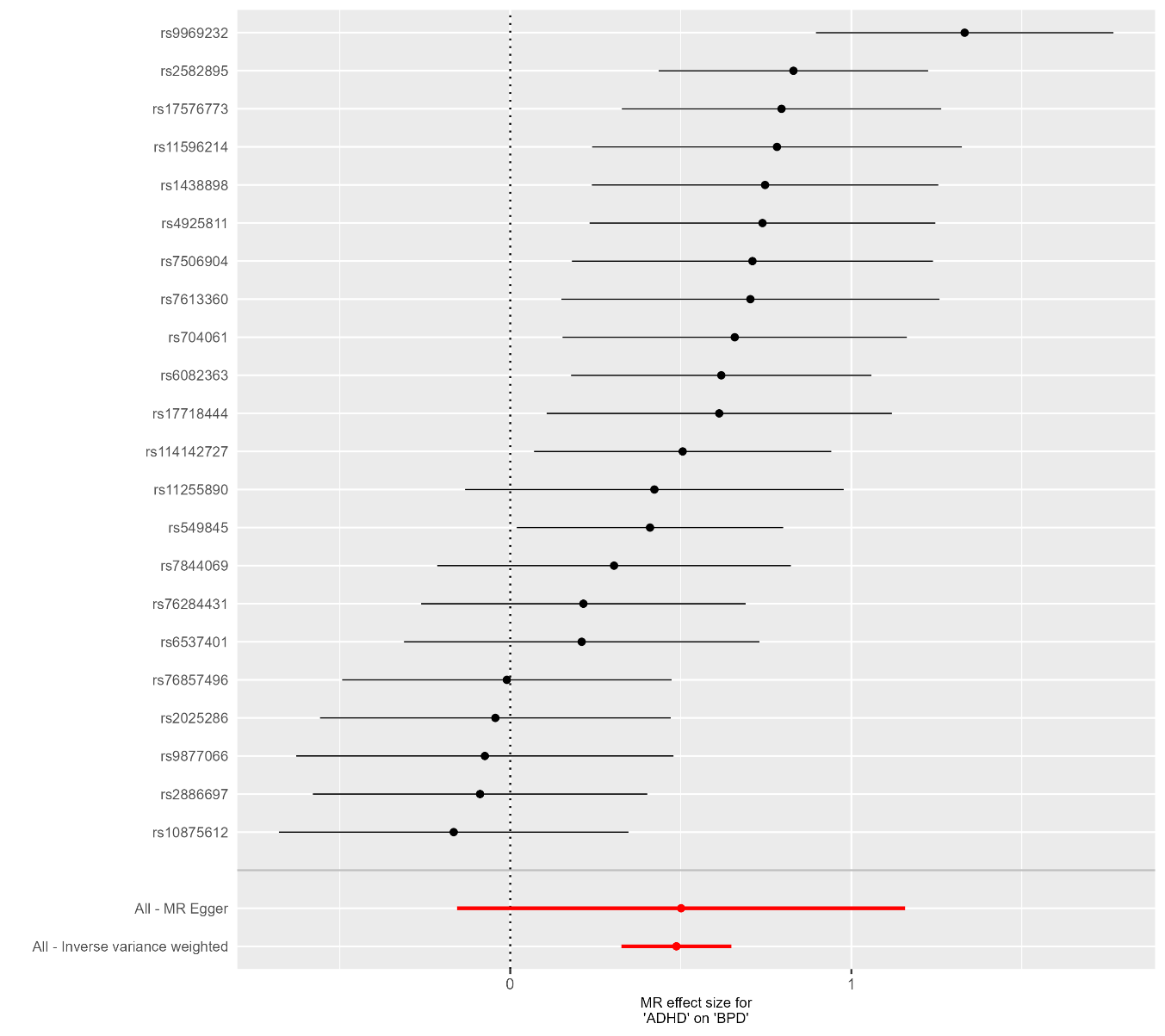

**Supplementary Figure 2. SNP-specific Mendelian randomization estimates for the association between genetic liability to attention-deficit/hyperactivity disorder and borderline personality disorder.** Individual points represent SNP-specific Wald ratio estimates. with horizontal lines indicating 95% confidence intervals. The red lines represents the overall inverse-variance weighted (IVW) and MR Egger estimates obtained using all 22 genetic instruments. The vertical dashed line indicates the null value.

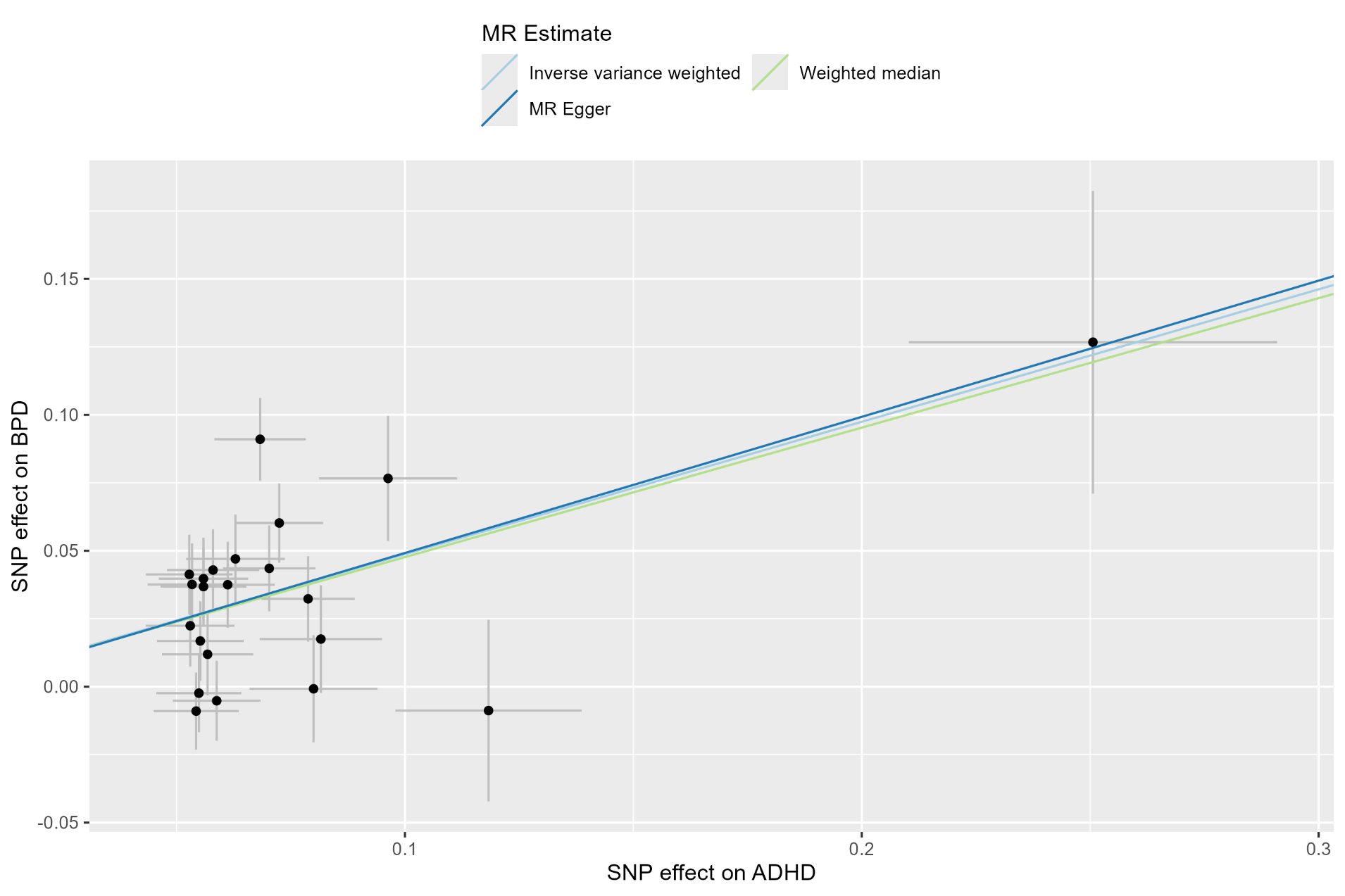

**Supplementary Figure 3**. MR-Egger scatter plot for the association of genetic liability to ADHD with BPD. Points represent individual genetic instruments. SNP effects on ADHD and BPD are plotted on the log-odds scale after orientation to the ADHD-increasing allele.

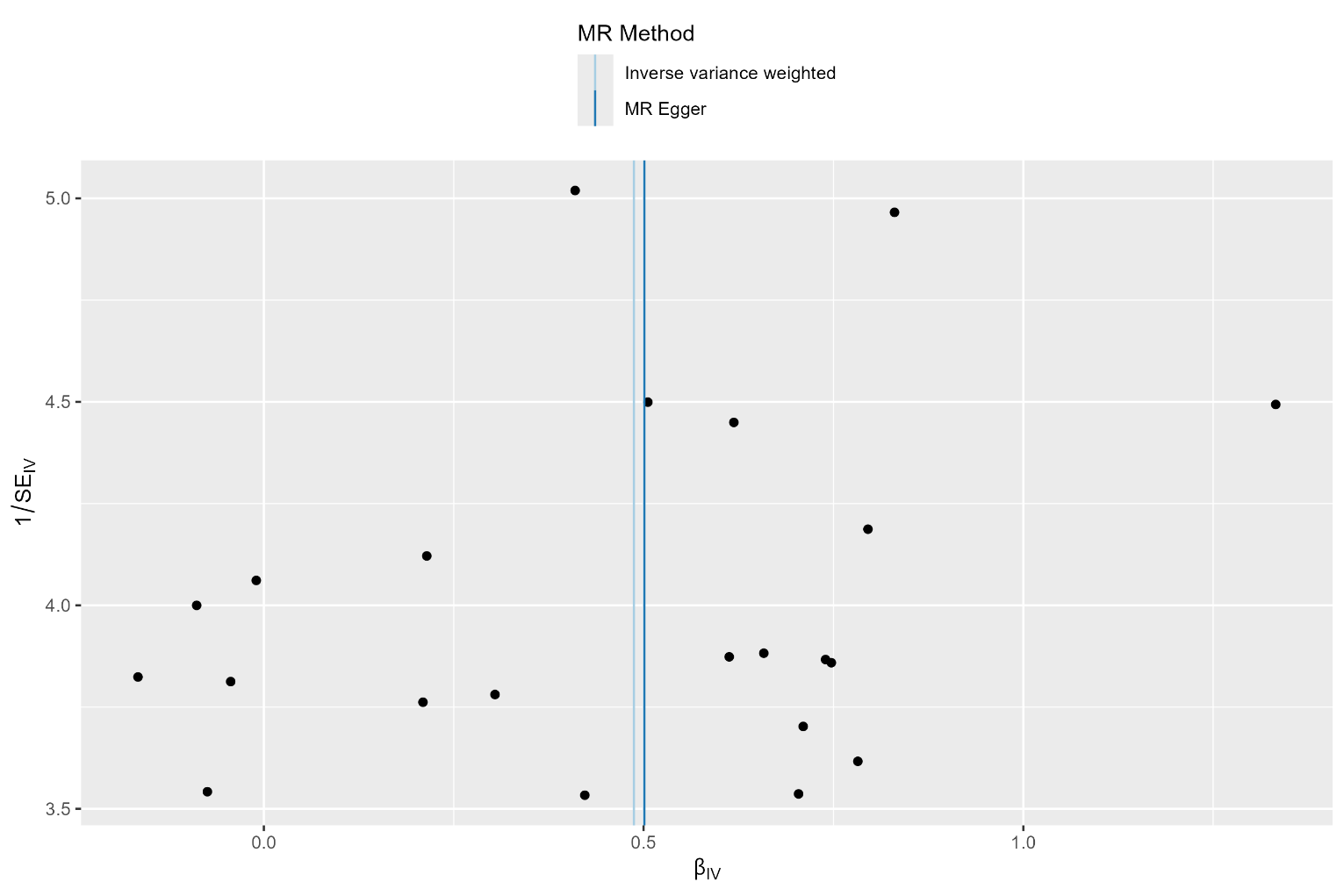

**Supplementary Figure 4** Funnel plot of Mendelian randomization estimates for the association between ADHD liability and borderline personality disorder (BPD). The plot displays SNP-specific Wald ratio estimates for the 22 independent genetic instruments. plotted against their inverse standard errors. Vertical lines indicate the pooled estimates obtained using inverse-variance weighted (IVW) and MR-Egger regression. The close agreement between the two estimates is consistent with their similar effect sizes. Formal assessment of directional horizontal pleiotropy using the MR-Egger intercept test was not statistically significant (p = 0.967). although significant between-instrument heterogeneity was observed (Cochran’s Q = 51.09; P = 2.57 × 10⁻⁴).

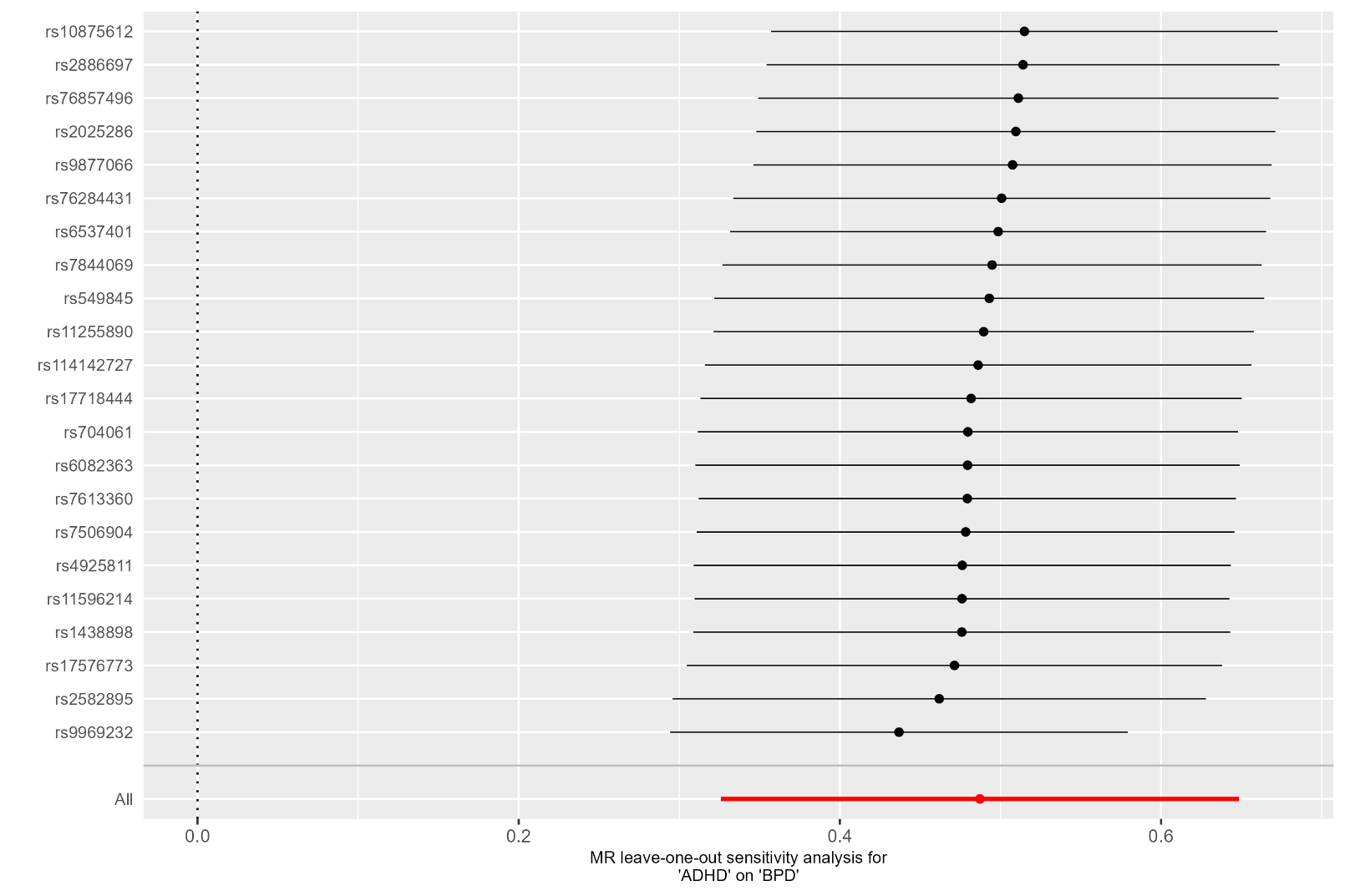

**Supplementary Figure 5**. Leave-one-out analysis of the association between genetic liability to attention-deficit/hyperactivity disorder and borderline personality disorder. Each point represents the inverse-variance weighted (IVW) Mendelian randomization estimate obtained after sequential exclusion of the indicated genetic instrument; horizontal lines represent 95% confidence intervals. The estimate obtained using all 22 genetic instruments is shown in red. The vertical dashed line indicates the null value. Estimates remained directionally consistent and statistically significant after exclusion of each individual instrument. supporting the robustness of the ADHD-to-BPD association to single-variant influence.

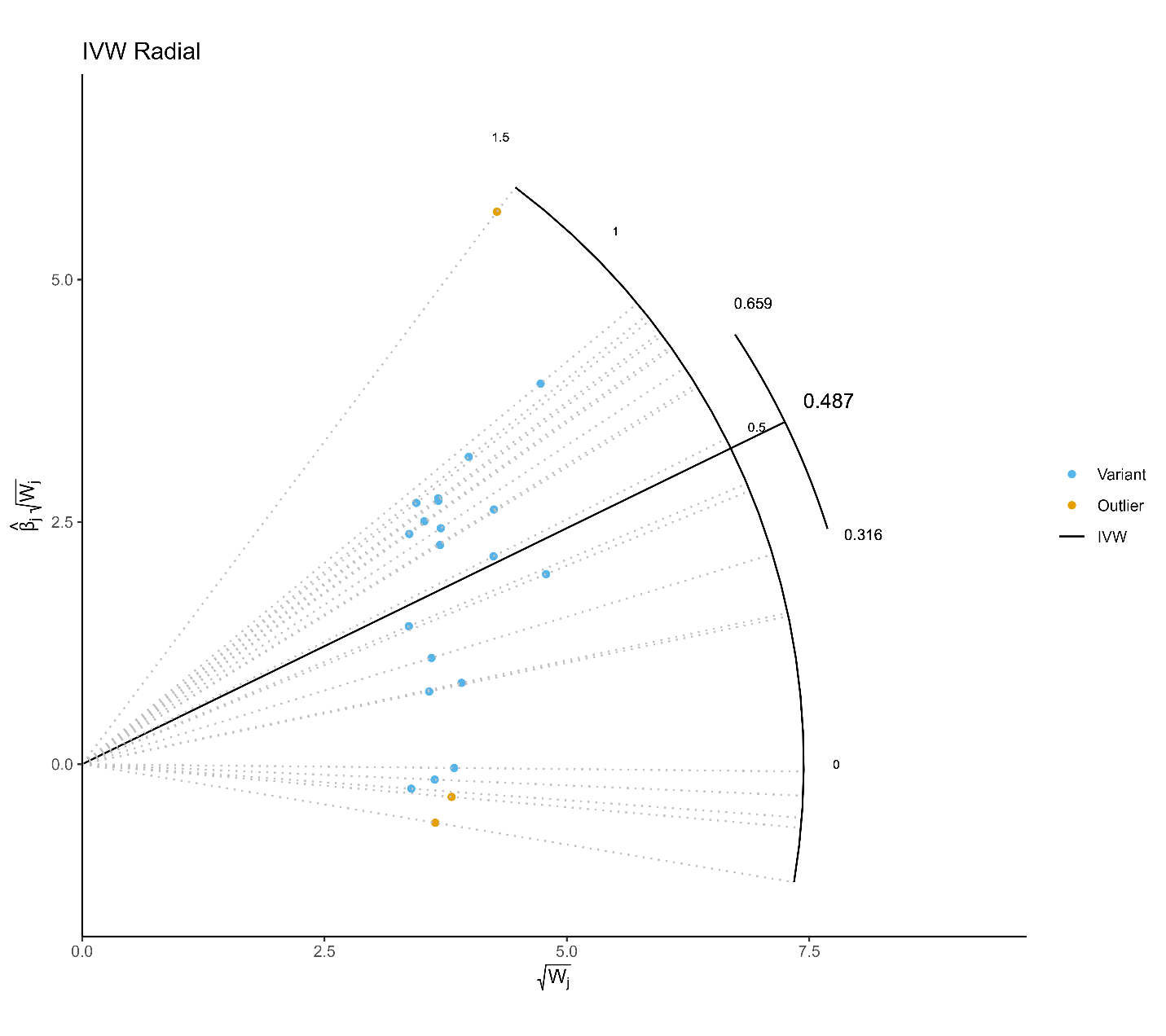

**Supplementary Figure 6. Radial Mendelian randomization plot for the association between genetic liability to attention-deficit/hyperactivity disorder and borderline personality disorder.** The radial plot displays the 22 genetic instruments included in the primary ADHD-to-BPD analysis using modified second-order weights. The solid line represents the Radial IVW estimate Variants highlighted as outliers showed an increased contribution to overall heterogeneity.

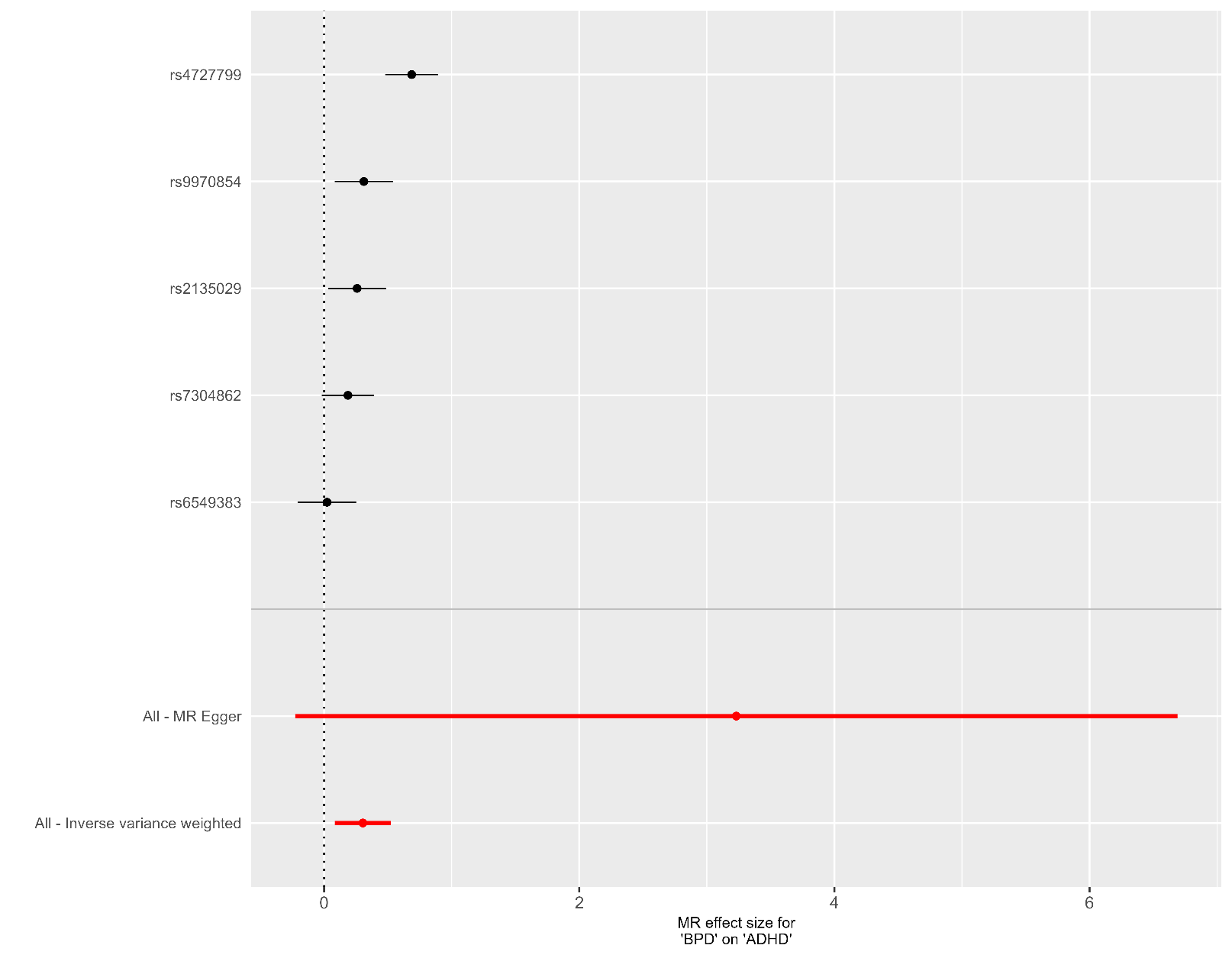

**Supplementary Figure 7. SNP-specific Mendelian randomization estimates for the association between genetic liability to borderline personality disorder and attention-deficit/hyperactivity disorder**. Individual points represent beta coefficents for attention-deficit/hyperactivity disorder derived from SNP-specific Wald ratio estimates. with horizontal lines indicating 95% confidence intervals. The red bars represents the overall inverse-variance weighted (IVW) and MR Egger estimates obtained using all five genetic instruments included in the analysis. The vertical dashed line indicates the null value.

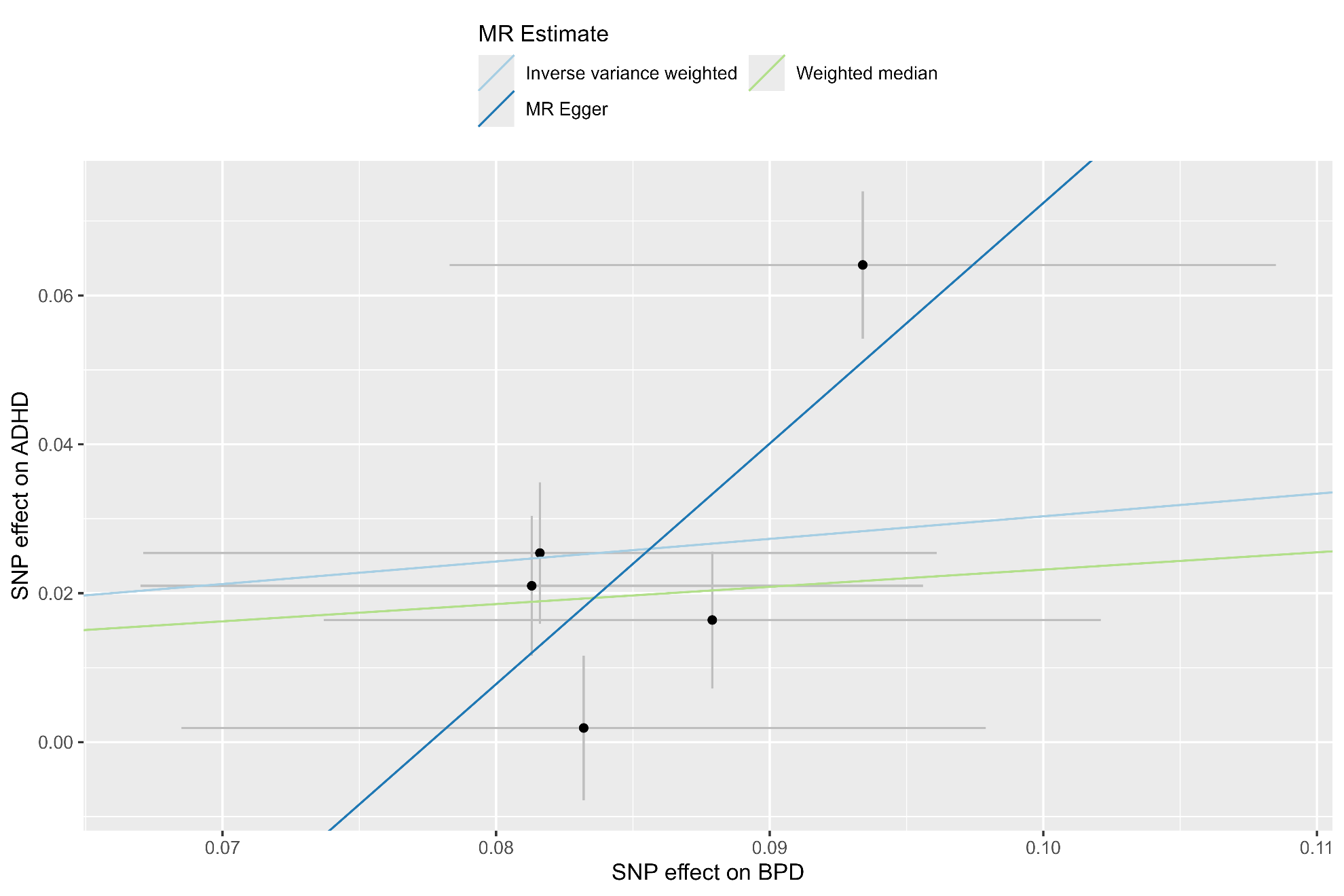

**Supplementary Figure 8. MR-Egger scatter plot for the association of genetic liability to BPD with ADHD.** Points represent individual genetic instruments. SNP effects on BPD and ADHD are plotted on the log-odds scale after orientation to the BPD-increasing allele. The dashed line represents the inverse-variance weighted estimate and the solid line the MR-Egger regression estimate. Although MR-Egger intercept did not provide statistically significant evidence of directional horizontal pleiotropy , this analysis was poorly informative because of the small number of genetic instruments

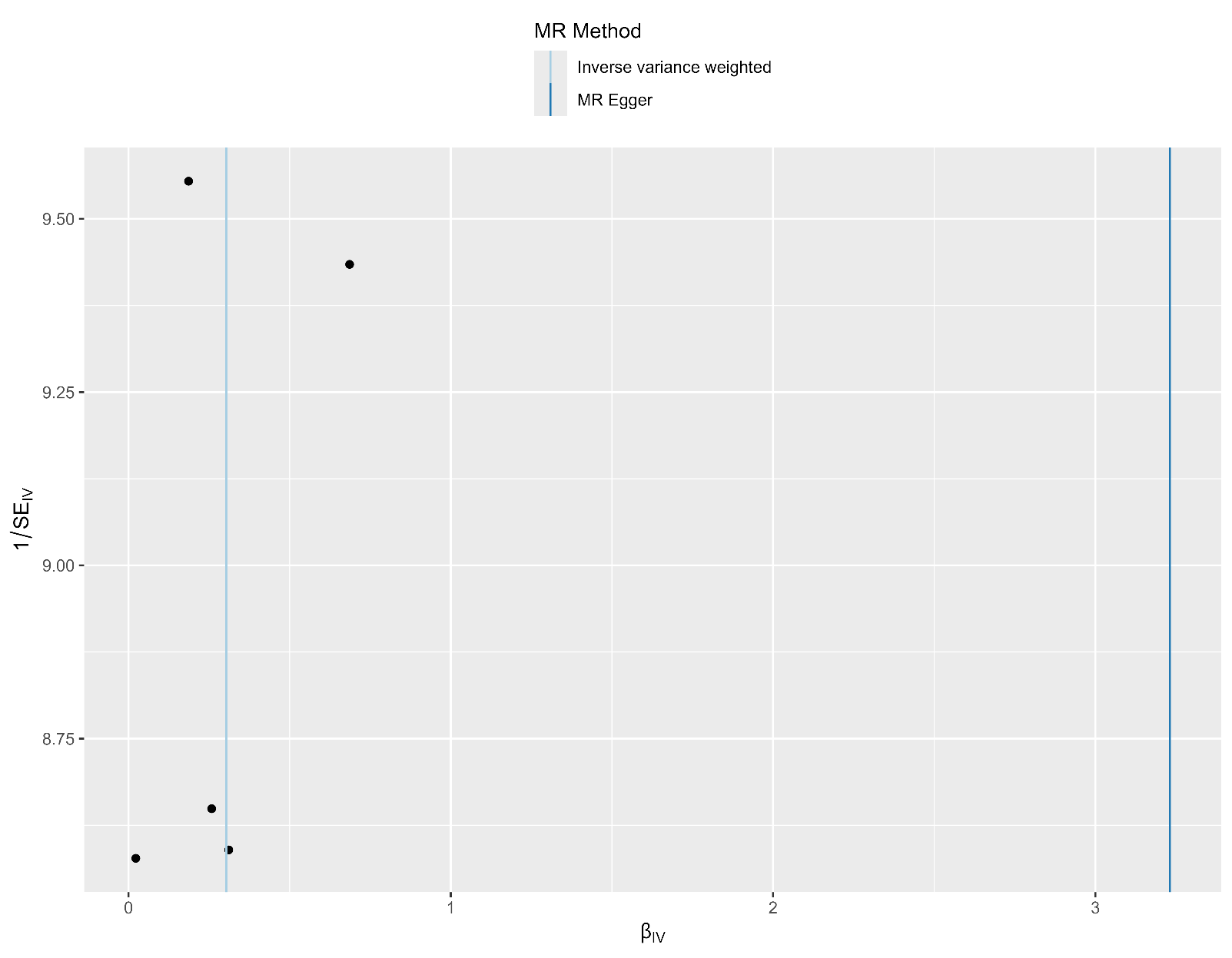

**Supplementary Figure 9. Funnel plot for the BPD-to-ADHD Mendelian randomization analysis.** Points represent SNP-specific Wald ratio estimates plotted against their precision (inverse standard error). Lines indicate the inverse-variance weighted and MR-Egger estimates. Interpretation of funnel-plot asymmetry is limited by the small number of instruments (n = 5).

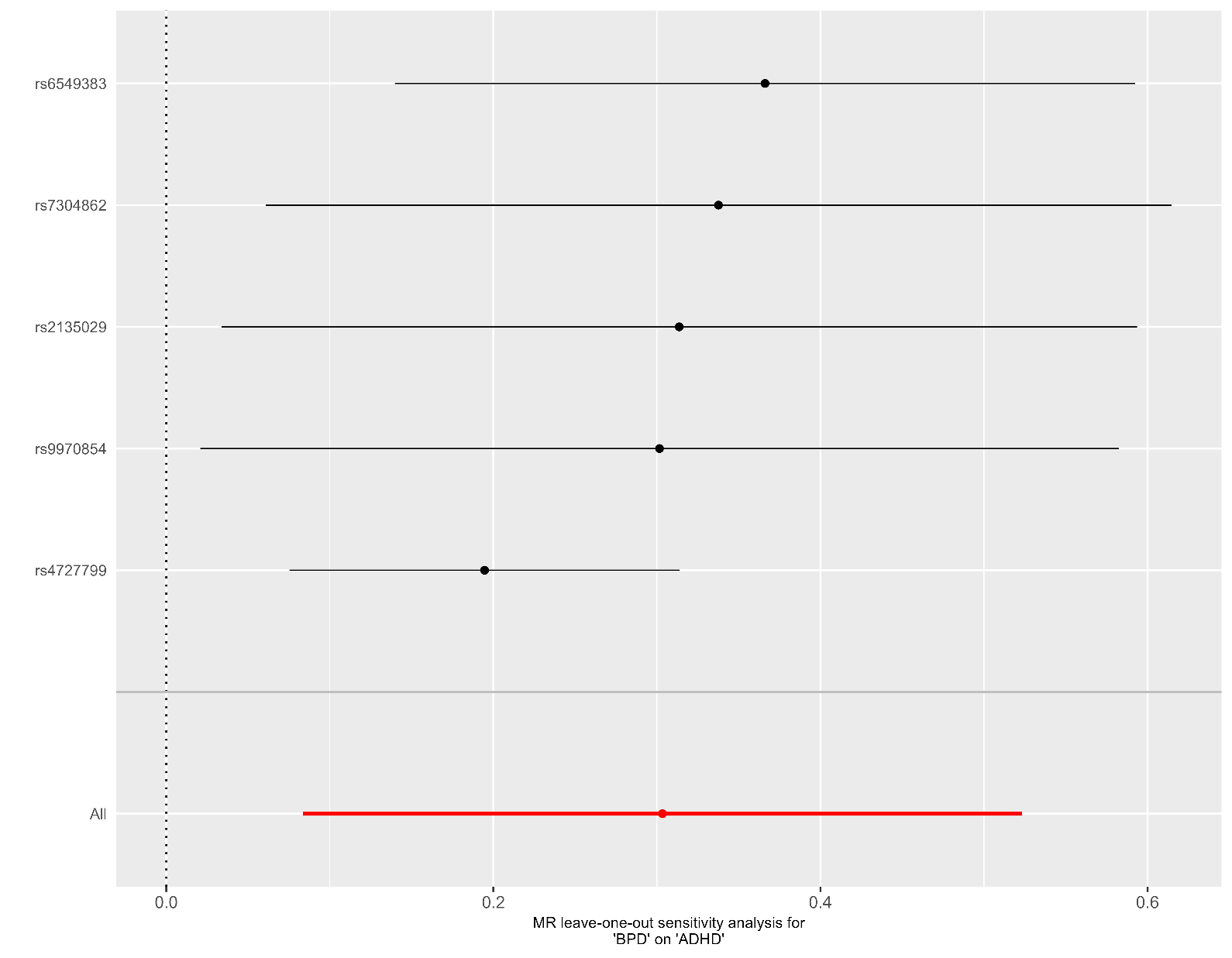

**Supplementary Figure 10. Leave-one-out analysis of the association between genetic liability to borderline personality disorder and attention-deficit/hyperactivity disorder.** Each point represents the inverse-variance weighted (IVW) Mendelian randomization estimate obtained after sequential exclusion of the indicated genetic instrument; horizontal lines represent 95% confidence intervals. The estimate obtained using all five genetic instruments is shown at the bottom. The vertical dashed line indicates the null value Estimates remained directionally consistent and statistically significant after exclusion of each individual instrument. Exclusion of rs4727799 reduced the effect estimate but preserved the association and substantially reduced between-instrument heterogeneity. indicating that this variant contributed to heterogeneity but did not account for the overall BPD-to-ADHD association.

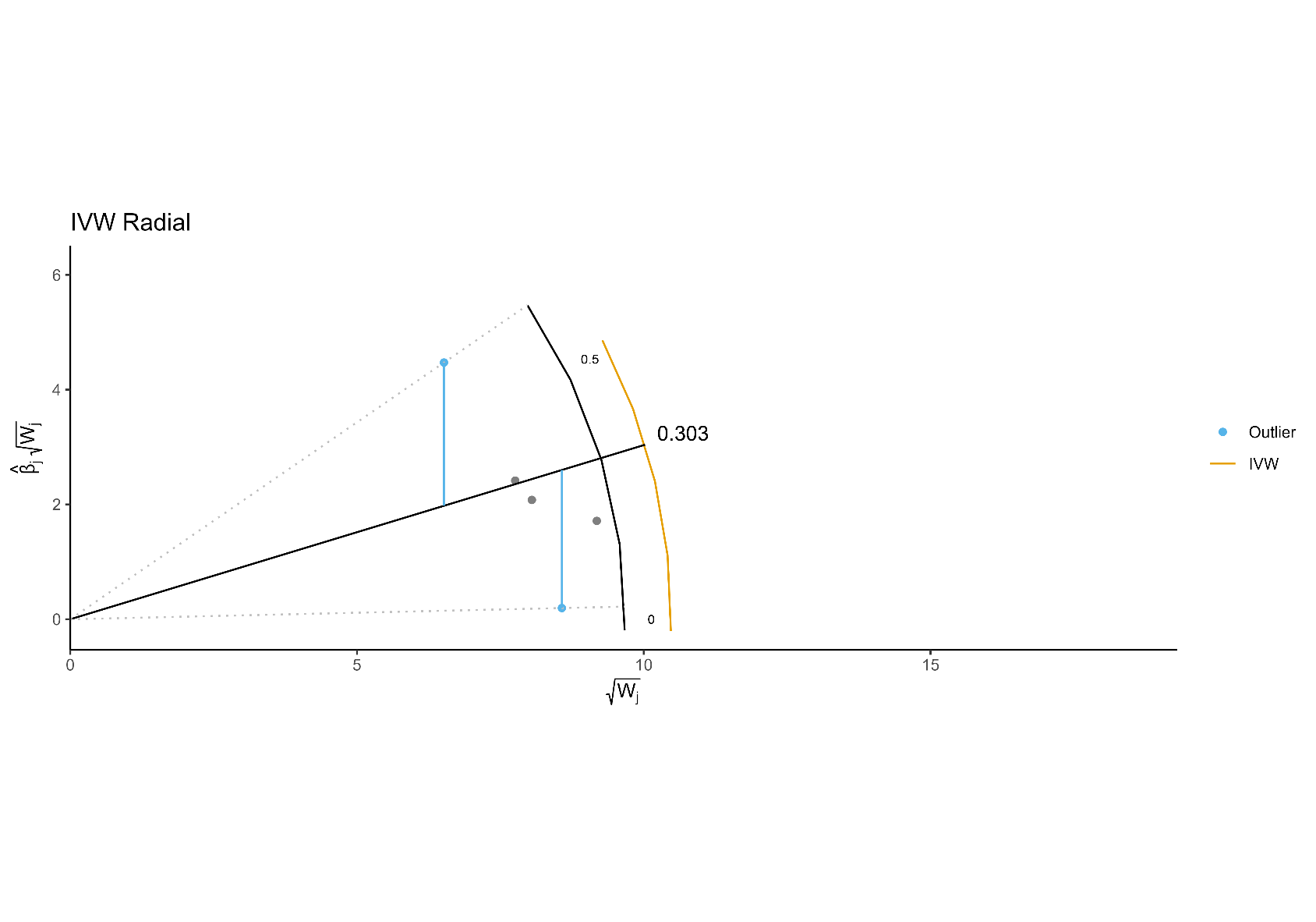

**Supplementary Figure 11. Radial Mendelian randomization plot for the association between genetic liability to borderline personality disorder and attention-deficit/hyperactivity disorder.** The radial plot displays the five genetic instruments included in the BPD-to-ADHD analysis using modified second-order weights. The solid line represents the Radial IVW estimate (β = 0.252. SE = 0.098; p = 0.0099). Two variants were nominally identified as radial outliers (rs6549383. p = 0.049. and rs4727799. p = 0.0047). although only rs4727799 remained significant after Bonferroni correction for five instruments (p < 0.01). Overall heterogeneity was significant (Q = 12.45. df = 4; p = 0.014).
